# Social contact patterns during the early COVID-19 pandemic in Norway: insights from a panel study, April to September 2020

**DOI:** 10.1101/2023.11.18.23298731

**Authors:** Lamprini Veneti, Bjarne Robberstad, Anneke Steens, Frode Forland, Brita A. Winje, Didrik F. Vestrheim, Christopher I Jarvis, Amy Gimma, W. John Edmunds, Kevin Van Zandvoort, Birgitte Freiesleben de Blasio

## Abstract

**Background:** During the COVID-19 pandemic, many countries adopted social distance measures and lockdowns of varying strictness. Social contact patterns are essential in driving the spread of respiratory infections, and country-specific measurements are needed. This study aimed to gain insights into changes in social contacts and behaviour during the early pandemic phase in Norway.

**Methods:** We conducted an online survey among a nationally representative sample of Norwegian adults, including six data collections/waves between April and September 2020, and used survey data from 2017 as baseline. We calculated mean daily contacts, and estimated age-stratified contact matrices that were used to estimate reproduction numbers during the study period.

**Results:** The mean daily number of contacts varied between 3.2 (95% CI 3.0-3.4) to 3.9 (95% CI 3.6-4.2) across waves, representing a 67-73% decline compared to pre-pandemic levels. Fewer contacts in the community setting largely drove the reduction; the drop was most prominent among younger adults. Despite gradual easing of social distance measures during the survey period, population contact matrices remained relatively stable and displayed more inter-age group mixing than at baseline. Contacts within households and the community outside schools and workplaces contributed most to social encounters.

**Conclusion:** Social contacts experienced a significant decline during the months following the March 2020 lockdown in Norway, aligning with the implementation of stringent social distancing measures. The findings contribute valuable empirical information into the social behaviour of the Norwegian population during the early pandemic, which can be used to enhance policy-relevant models for addressing future crises when mitigation measures might be implemented.

## Introduction

During the beginning of 2020, COVID-19 rapidly spread around the globe, resulting in a considerable burden on public health and economic welfare of societies (1). In order to limit the burden of the disease and prevent a collapse of healthcare services, governments early imposed regulations requiring people to reduce social interactions and other risky behaviours in an effort to contain the transmission of COVID-19 (2, 3).

In Norway, the first COVID-19 case was detected on 26 February (4). Following a rapid increase in COVID-19 hospitalisations, on 12 March, the Norwegian government issued a national lockdown with strict border control, mandatory home quarantine and isolation in case of exposure and infection, closure of educational institutions and shops except for essential goods and medicine and cancellation of sports and cultural events. In addition, there were recommendations to increase hygiene, work from home if possible, and avoid public transportation and domestic travel (4, 5). After a subsequent drop in COVID-19 hospitalisations, a gradual reopening started late April 2020, prioritising day-care and primary schools, later followed by high schools, and allowance of public events of limited size (4, 5). In June 2020, bars and sports activities could reopen under 1-metre social distance conditions, and international travel restrictions were scaled back, except for regions with high infection levels. COVID-19 hospitalisations remained low in Norway throughout the summer and until mid-autumn. By October 2020, it was estimated that 90 000, corresponding to 1.6% of the population had been infected (6).

Mathematical models were key in informing public health policy decision-making during the COVID-19 pandemic. The models provided a framework for informed assessment of the epidemic evolution, short-term forecasting, and estimation of effects of control measures based on transparent assumptions and data. However, the models were constrained by scarce knowledge about the virus, infection prevalence, immune response, uncertainties related to data, including underreporting of cases, and, not least, missing data about social mixing patterns during a period of unprecedented control measures (7). Due to the dynamic nature of the COVID-19 epidemic, the models needed constant updates to provide timely, data-driven estimates of the effective reproduction number (8, 9) and short-term projections of the health-care burden (10).

SARS-CoV-2 is primarily transmitted via respiratory droplets and close contact routes with use of quantitative country- and age-specific data on social mixing has been found essential in models to study transmission dynamics of close-contact infections (11–13). In early 2020, social contact pattern data for Norway were available from a survey conducted in 2017 (14), inspired by the earlier POLYMOD survey (12). These social mixing pattern data collected during “peace time” were unlikely to be valid during the pandemic. Instead, the behaviour was expected to change during the rapidly evolving crisis in response to policies, disease incidence and health risk perceptions, and there was an urgent need for updated social mixing data to support the ongoing management of the disease and improve the general understanding of contact patterns of relevance for outbreak preparedness.

Here, we report on a panel study conducted in Norway during the first year of the pandemic, providing novel information about social mixing patterns during the early phase of the pandemic. By comparing the survey results to Norwegian data collected in a 2017 survey, we aimed to quantify changes in the social contacts and mixing patterns and, consequently, their impact on the transmissibility of SARS-CoV-2.

## Methods

### Data collection and recruitment

The Norwegian panel survey was part of the international online CoMix study collecting data on social contact patterns, behaviour and attitudes over the course of the COVID-19 pandemic in European countries (15). The CoMix study was inspired by the POLYMOD study (12) and was launched in March 2020 in United Kingdom (UK), Belgium and the Netherlands. The Norwegian Institute of Public Health and the University of Bergen joined this collaboration, resulting in a 6-wave Norwegian panel survey conducted between April and October 2020. Details on the CoMix study, including the protocol and survey questionnaire have been published previously (16). We translated the CoMix survey questionnaire into Norwegian.

The market research company Ipsos was responsible for carrying out the survey in Norway and other countries participating in the CoMix study. In Norway, Ipsos recruited a nationally representative adult population sample with respect to age, gender, geographical location, and socio-economic status from an existing online panel (17). The company developed the web-based form from the translated CoMix questionnaire.

In each of the six waves, participants were invited by email to register their contacts on a randomly assigned weekday of each week of data collection. In case of non-response, up to two reminder emails were sent. To account for dropouts, Ipsos recruited additional panellists during the study period with similar characteristics in terms of age and gender. Participants received “panellist points” exchangeable for shopping vouchers for each questionnaire they filled out. Fig. 1 shows an overview of the Norwegian CoMix survey with dates and numbers of participants and substitutes recruited in the waves.

**Fig. 1:**
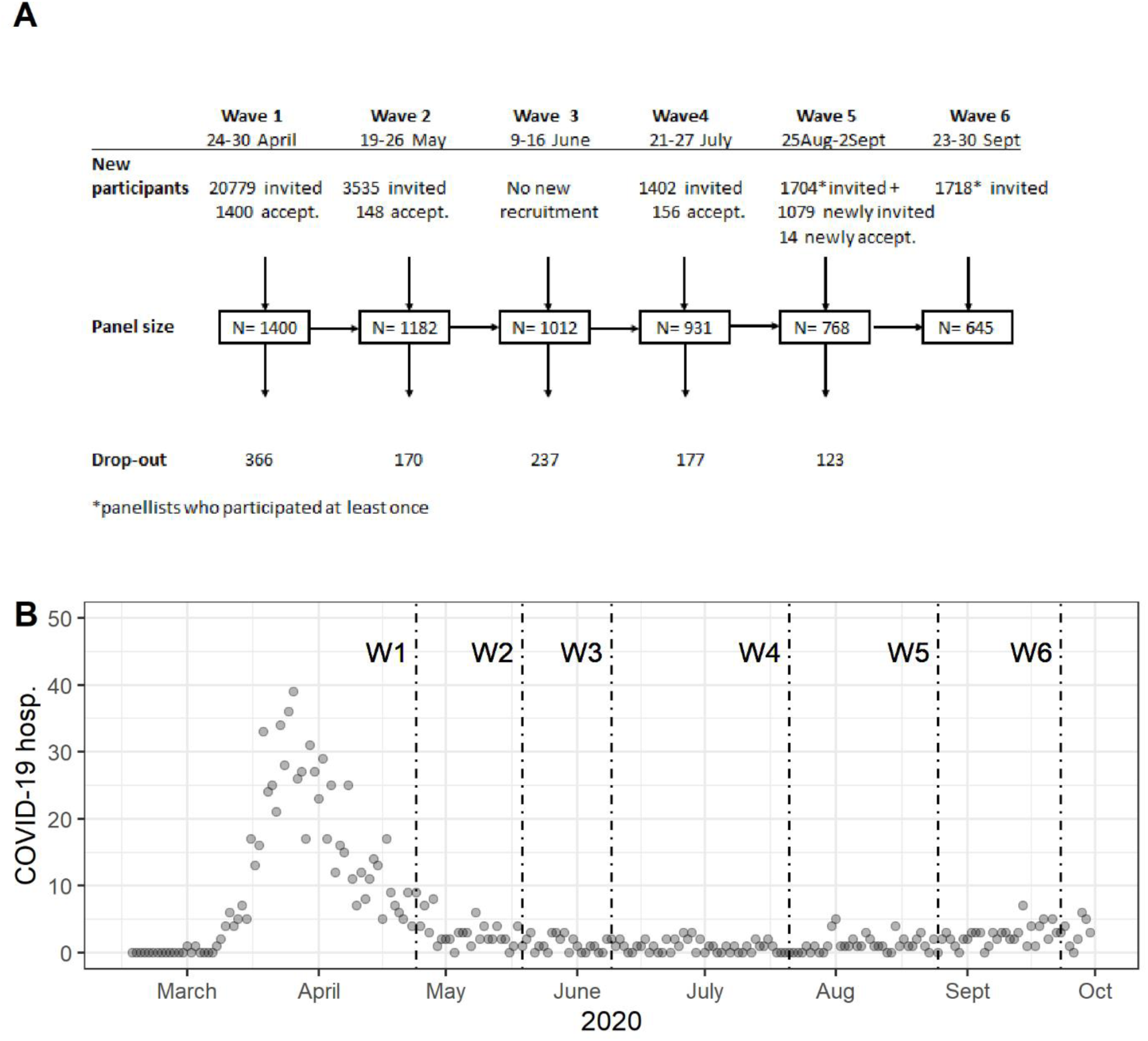
Norwegian CoMix survey: (A) Overview of recruitment and data collection, April to September 2020, (B) shows the number of daily hospital admissions in Norway during the survey period, with start dates for each wave indicated with a vertical line.

Participants provided socio-economic and demographic information during the enrolment survey. Other data were collected using an identical questionnaire in each wave. The questionnaire included detailed questions about their social contacts, exposure to social distancing measures, uptake of preventive measures, and attitudes and risk perception regarding the COVID-19 pandemic. This article focuses on the social contact data and related personal protection measures (hand washing, use of hand sanitiser and face masks) while analyses of other survey topics have been published previously (17, 18) or are planned to be published soon.

### Social contact data

CoMix survey participants were asked to detail their social contacts on the day preceding the email invitation (24 hours). The questionnaire defined a social contact as either an in-person conversation (exchange of at least few words) or physical contact (involving touch, e.g., handshake, embracing, contact sports). Participants could register up to 45 individual contacts per survey wave with information about the (estimated) age and gender of the contact, their relations, location of the encounter (home, work, school etc.), duration and frequency of interaction, type of contact (physical or not) and whether the contact occurred outdoors or indoors. In addition, questions on group contacts followed the questions on individual contacts and were added to surveys after wave 1. Group contacts were added to accommodate participants who could not record details of all individual contacts that they had, e.g., clients, patients, students etc. To address potential biases, uncertainties, and disparities between the CoMix and baseline survey questionnaires, we made the decision to exclude grouped contacts from our current study. There were several reasons behind this choice. Firstly, the first CoMix wave did not gather grouped data, making it impossible to ensure comparability of contact numbers across different waves. Secondly, we lacked information regarding the age distribution of grouped contacts in the 2017 baseline survey. Lastly, we identified instances where a few individuals reported exceptionally high contact numbers, which, in certain cases, were due to typographical errors or misunderstandings. More details on group contacts are presented in Online Resource, Section 1.

### Baseline survey

We compared the Norwegian CoMix data to the Norwegian social contact data collected through a cross-sectional survey (single wave) conducted by the Norwegian Institute of Public Health in 2017 (14). Participants were randomly selected from the population registry by Statistics Norway (19) and received invitations and paper questionnaires by mail. Enrolment occurred in late April with a reminder sent to non-responding invitees in September. In the study 4 792 people were invited, of which 2 593 were adults (≥18 years). From the invitees, 565 (response rate, 12%) individuals aged 0 to 92 years participated including 309 (response rate, 12%) adults participate.

Participants were asked to fill in their social contacts on a particular weekday using a paper diary similar to the one used in the POLYMOD study (12). They could report up to 49 individual contacts, and additional grouped contacts described in free text format (group contacts were excluded as explained above). Because the 2017 Norwegian questionnaire closely resembles the CoMix survey, we used it as baseline measurement of social contacts during “peace time” (pre-pandemic) in Norway. Henceforth, the study is referred to as the ‘baseline survey’.

### Data analysis

We compared the age, gender, and county of residence of our survey participants to the 2020 mid-year estimates of the Norwegian adult population provided by Statistics Norway (19) to assess the representativeness of our study population in the different waves.

We computed the mean number of daily contacts for each CoMix wave, stratified by participant characteristics, including age group, gender, household size and day of the week. We also present comparative results with the baseline survey as reference. Using the survey design in Stata 16.0, for each wave, we weighted the analysis by age group and gender to obtain population-representative results. The weighted mean number of contacts were presented with 95% confidence intervals (CI). Data on personal protective measures were analysed by calculating percentages of participants reporting hand washing and using hand sanitisers and face masks.

We calculated age-specific contact matrices for both the CoMix waves and the baseline survey, categorising age groups into 0-4, 5-17, 18-29, 30-49, 50-69, and 70+ years. According to the social contact hypothesis, we assumed that the age-specific number of social contacts is directly proportional to the potential transmission events (11). In this context, the next generation transmission matrix, denoted as *NG*, is linked to the population contact matrix as NG= (*ng_ij) = q* where *q* represents the infectivity factor, considering infectivity and the duration of the infectious period. Each element of the next generation matrix, *ng_ij*, represents the anticipated number of new infections in age group *i* caused by an infected individual in age group *j*, under the assumption of a completely susceptible population. The basic reproduction number, R0, is obtained as the largest eigenvalue of this matrix. In this analysis, we employed the social contact hypothesis to gauge shifts in transmissibility owing to changes in contact pattern during the early stages of the pandemic. This was achieved by determining the ratio between the largest eigenvalues of the CoMix contact matrices and that obtained from the 2017 survey.

Given that no data were collected for children under 18 years in the CoMix study, we employed imputation to generate complete population matrices (20). Initially, we conducted bootstrapping with replacement, creating 10 000 adult-to-adult matrices from the 2017 baseline study and each CoMix wave. We adjusted these matrices for the reciprocity of contacts between age groups and the mid-year age populations data from Statistics Norway (19). For each pair of samples, we calculated a scaling factor, denoted *f,* which was derived from the ratio of the maximum eigenvalues (*max λ*_CoMix / *max λ*_baseline), see Online Resource, Figure S3.

To estimate children-to-children contacts (0–4,5–17), we multiplied the average scaling factor by 10 000 bootstrapped full baseline population matrices, adjusting for the reciprocity of contacts and age to enhance representativeness, using the 2017 mid-year population data. Children-to-adult contacts were determined based on the adult-to-children contacts in the CoMix data, employing a similar bootstrapping approach with 10 000 full data matrices and age adjustments. Finally, the imputed matrices were further adjusted for reciprocity of contacts among age groups to match the 2020 mid-year population. This entire process was conducted separately for all contacts and physical contacts, and for different locations (home, school, work, and ’other’).

We compared the imputed population matrices of the CoMix waves and their dominant eigenvalues and dominant eigenvectors against those of the baseline survey. Additionally, we characterised the social mixing pattern by calculating the disassortativity index, as suggested by Farrington et al. (21). The index was standardized relative to homogeneous mixing, with a value of 1 indicating such mixing. Values greater than 1 indicated disassortative mixing with a preference for contacts outside one’s own age group, while values less than 1 indicated assortative mixing with a tendency for contacts within one’s own age group.

To estimate the early COVID-19 reproduction numbers in Norway, we scaled a model-derived estimate of R0 before the lockdown (10, 22), by the ratio of matrix eigenvalues (Imputed CoMix/baseline) for each wave and type of contact. In the 2017 baseline survey we performed multivariate imputation by chained equations (N=10 datasets) to account for missing values of physical contacts (10.8%), age of contacts (1.7%) and location (0.4%) using the R-package MICE. When adjusting for age, we used a threshold value of 3 to limit the influence of single participants.

We used Stata version 16 (Stata Corporation, College Station, Texas, US) and R version 4.0.0 with the socialmixr package ver. 0.2.0 and the MICE package ver. 3.15.0 to analyse the data.

## Results

### Sample characteristics

The Norwegian CoMix study included 1 718 participants and 5 938 reported questionnaires with information about a total of 22 074 contacts. In the first wave, 20 779 adults were invited, and 1 400 participated. Despite further recruitment, the sample size gradually decreased during subsequent waves reaching 645 participants in the final wave (Fig 1, panel A). Of the original 1 400 participants, 315 took part in all six waves.

The demographic characteristics of the participants in the CoMix waves and adults in the baseline survey are presented in Table 1. Approximately half of the participants were male, and the median age varied between 49 to 54 years across the waves, compared to 47 years in the Norwegian adult population. Additionally, the median age showed a slight increase over the study period. The mean household size was 2 with a range from 1 to 12. While panellists from all counties in Norway participated, Oslo was somewhat over-represented in all waves. Further details regarding the representativeness are provided in Online Resource, Section 2.

**Table 1.** Descriptive characteristics in the 2017 baseline survey and in each wave of the CoMix study in 2020 (starting date of the data collection is indicated in brackets).

| Characteristics | Number of participants (%) |  |  |  |  |  |  |
| --- | --- | --- | --- | --- | --- | --- | --- |
|  | Baseline survey 2017<br>n=309 | Wave 1<br>(24 April),<br>n=1400 | Wave 2<br>(19 May),<br>n=1182 | Wave 3<br>(9 June),<br>n=1012 | Wave 4<br>(21 July),<br>n=931 | Wave 5<br>(25 Aug),<br>n=768 | Wave 6<br>(23 Sept),<br>n=645 |
| **Gender** |  |  |  |  |  |  |  |
| Male | 146 (47 %) | 702 (50 %) | 610 (52 %) | 534 (53 %) | 487 (52 %) | 421 (55 %) | 357 (55 %) |
| Female | 163 (53 %) | 698 (50 %) | 569 (48 %) | 475 (47 %) | 441 (47 %) | 345 (45 %) | 287 (44 %) |
| Missing | 0 (0 %) | 0 (0 %) | 3 (0 %) | 3 (0 %) | 3 (0 %) | 2 (0 %) | 12 (1 %) |
| **Age group** |  |  |  |  |  |  |  |
| 18-29 | 46 (15 %) | 217 (16 %) | 121 (10 %) | 98 (10 %) | 78 (8 %) | 57 (7 %) | 70 (11 %) |
| 30-49 | 36 (12 %) | 509 (36 %) | 410 (35 %) | 352 (35 %) | 300 (32 %) | 257 (34 %) | 210 (33 %) |
| 50-69 | 93 (30 %) | 493 (35 %) | 458 (39 %) | 406 (40 %) | 406 (44 %) | 319 (42 %) | 262 (41 %) |
| 70+ | 134 (43 %) | 181 (13 %) | 193 (16 %) | 156 (15 %) | 147 (16 %) | 135 (18 %) | 103 (16 %) |
| **Day of week** |  |  |  |  |  |  |  |
| Weekday | 221 (72 %) | 1003 (72 %) | 955 (81 %) | 930 (82 %) | 782 (84 %) | 584 (76 %) | 630 (98 %) |
| Weekend | 87 (28 %) | 397 (28 %) | 227 (19 %) | 82 (18 %) | 149 (16 %) | 184 (24 %) | 15 (2 %) |
| Missing | 1 (0 %) | 0 (0 %) | 0 (0 %) | 0 (0 %) | 0 (0 %) | 0 (0 %) | 0 (0 %) |
| **Household size** |  |  |  |  |  |  |  |
| 1 | 64 (21 %) | 320 (23 %) | 300 (25 %) | 278 (27 %) | 269 (29 %) | 200 (26 %) | 179 (28 %) |
| 2 | 169 (55 %) | 453 (32 %) | 458 (39 %) | 387 (38 %) | 358 (38 %) | 315 (41 %) | 251 (39 %) |
| 3-4 | 53 (17 %) | 482 (35 %) | 333 (28 %) | 275 (27 %) | 258 (28 %) | 216 (28 %) | 177 (27 %) |
| 5+ | 16 (5 %) | 145 (10 %) | 91 (8 %) | 72 (7 %) | 46 (5 %) | 37 (5 %) | 38 (6 %) |
| Missing | 7 (2 %) | 0 (0 %) | 0 (0 %) | 0 (0 %) | 0 (0 %) | 0 (0 %) | 0 (0 %) |
| **Education** |  |  |  |  |  |  |  |
| Nursery, primary school | 32 (10 %) | 57 (4 %) | 42 (4 %) | 39 (4 %) | 41 (5 %) | 28 (4 %) | 26 (4 %) |
| Secondary school | 121 (39 %) | 349 (25 %) | 282 (24 %) | 232 (23 %) | 216 (23 %) | 186 (24 %) | 153 (24 %) |
| Higher education<sup>b</sup> | 151 (49 %) | 994 (71 %) | 858 (72 %) | 741 (73 %) | 674 (72%) | 554 (72 %) | 466 (72 %) |
| Missing or Other | 5 (2 %) | 0 (0 %) | 0 (0%) | 0 (0 %) | 0 (0 %) | 0 (0 %) | 0 (0 %) |
| **Daily activity** |  |  |  |  |  |  |  |
| At home<sup>a</sup> | 188 (61 %) | 437 (31 %) | 416 (35 %) | 356 (35 %) | 371 (40 %) | 392 (51 %) | 238 (37 %) |
| School/<br>education | 28 (9 %) | 110 (8 %) | 63 (5 %) | 49 (5 %) | 34 (4 %) | 355 (46 %) | 35 (5 %) |
| Employed | 77 (25 %) | 853 (61 %) | 703 (60 %) | 607 (60 %) | 526 (57 %) | 21 (3 %) | 372 (58 %) |
| Missing or Other | 16 (3 %) | 0 (0 %) | 0 (0 %) | 0 (0 %) | 0 (0 %) | 0 (0 %) | 0 (0 %) |
| **High risk group<sup>c</sup>** |  |  |  |  |  |  |  |
| No | NA | 791 (57 %) | 641 (54 %) | 560 (55 %) | 472 (51 %) | 392 (51 %) | 351 (54 %) |
| Yes | NA | 561 (40 %) | 510 (43 %) | 425 (42 %) | 433 (46 %) | 355 (46 %) | 275 (43 %) |
| Do not know/ do not wish<br>to answer | NA | 48 (3 %) | 31 (3 %) | 27 (3 %) | 26 (3 %) | 21 (3 %) | 19 (3 %) |
| **Hand hygiene (washed**<br>**hands/ used sanitizer)<sup>d</sup>** |  |  |  |  |  |  |  |
| No | NA | 63 (5 %) | 65 (6 %) | 58 (6 %) | 44 (5 %) | 39 (5 %) | 24 (4 %) |
| Yes | NA | 1 337 (95 %) | 1 117 (94 %) | 954 (94 %) | 887 (95 %) | 729 (95 %) | 621 (96 %) |
| **Used Face Mask<sup>d</sup>** |  |  |  |  |  |  |  |
| No | NA | 1 339 (96 %) | 1 135 (96 %) | 981 (97 %) | 907 (97 %) | 719 (94 %) | 585 (91 %) |
| Yes | NA | 61 (4 %) | 47 (4 %) | 31 (3 %) | 24 (3 %) | 49 (6 %) | 60 (9 %) |

NA: not available
a: In this category we have included people who reported they have retired, were unable to work.
b: As higher education, we have grouped together people that had “Occupational training/Technical school”, “Higher education <4 years” and “Higher education ≥4 years”.
c: Risk groups were defined as the one that are recommended to be vaccinated for influenza in UK. https://www.nhs.uk/conditions/vaccinations/who-should-have-flu-vaccine/. Defined on the question “Are you in a high-risk group under which the annual influenza vaccine would usually be offered?”. Originally 516 participants reported that they had high risk of COVID-19 infection or complications, but after assessing the available information (including pregnant women and people above 65 years old) we assigned 561 (40%) to belong in a risk group. Overall, few pregnant women participated in the Comix study ranging from 4 to 19 over the six waves (around 1% per wave).
d: In the CoMix survey, participants reported whether they had at least once washed their hands with soap or used hand sanitiser during the last three hours prior to the survey. Regarding the use of face mask, participants were asked whether they used a face mask during the day prior to the survey day.

### Drop in the mean number of contacts

In the baseline survey, 2.3% of the participants reported zero contacts on the survey day, while in the CoMix study this was 8% (533/5 938) overall. This number increased from 6% in April 2020 to 9-12% in the following waves.

The mean number of daily contacts remained low and stable between April and September, varying from 3.2 (95% CI 3.0 –3.4) in July (wave 4) to 3.9 (95% CI 3.6–4.2) in June (wave 3) (Table 2). In comparison, in the baseline survey the mean number of contacts among the adults was 11.9 (95% CI 10.2-13.5), indicating a striking 67-73% decline in social contacts. The largest drop was found in younger adults aged 18-49 years, while in people aged 70 years and older the number was affected least (Figure 2A).

**Table 2:**
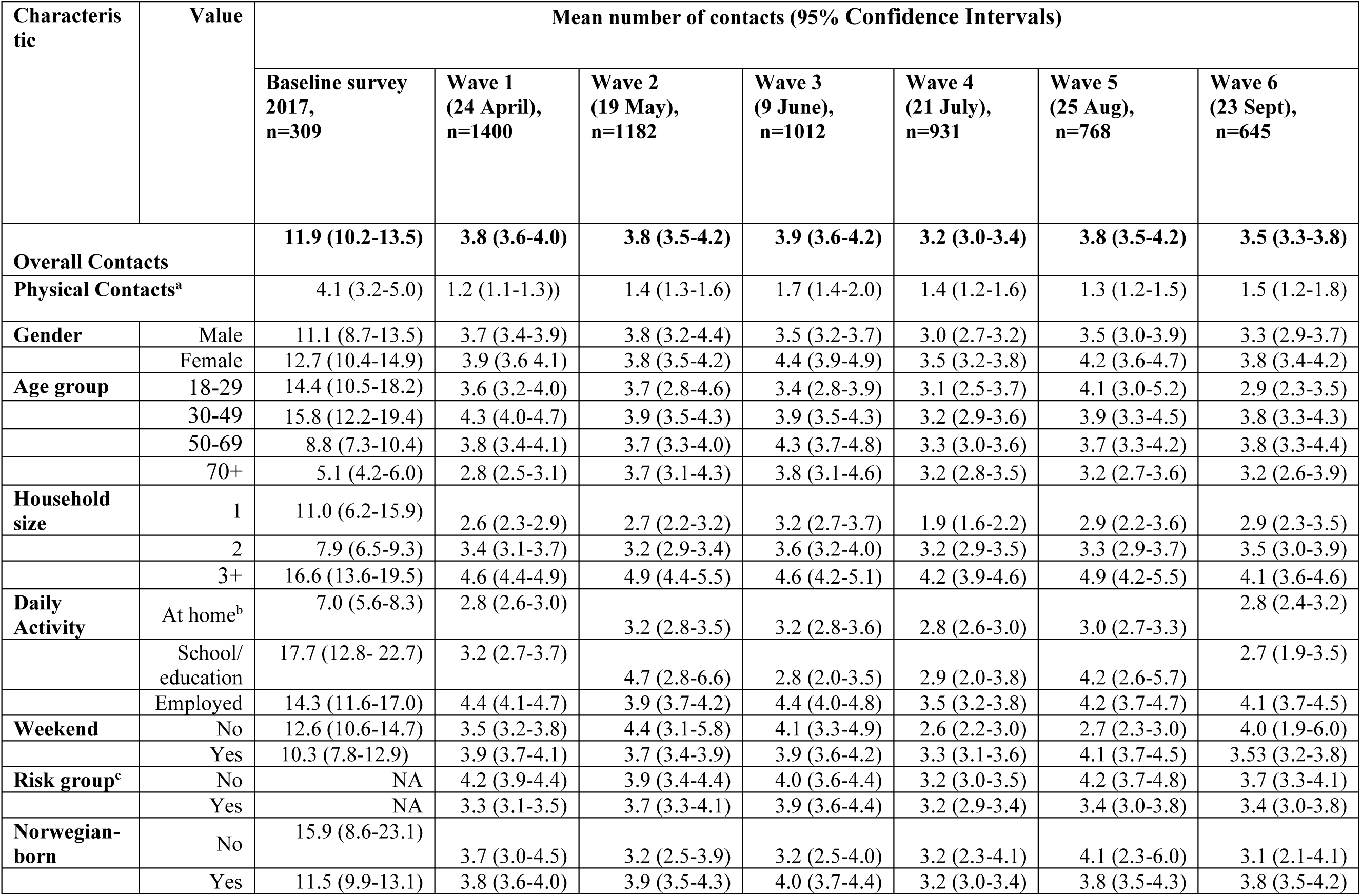
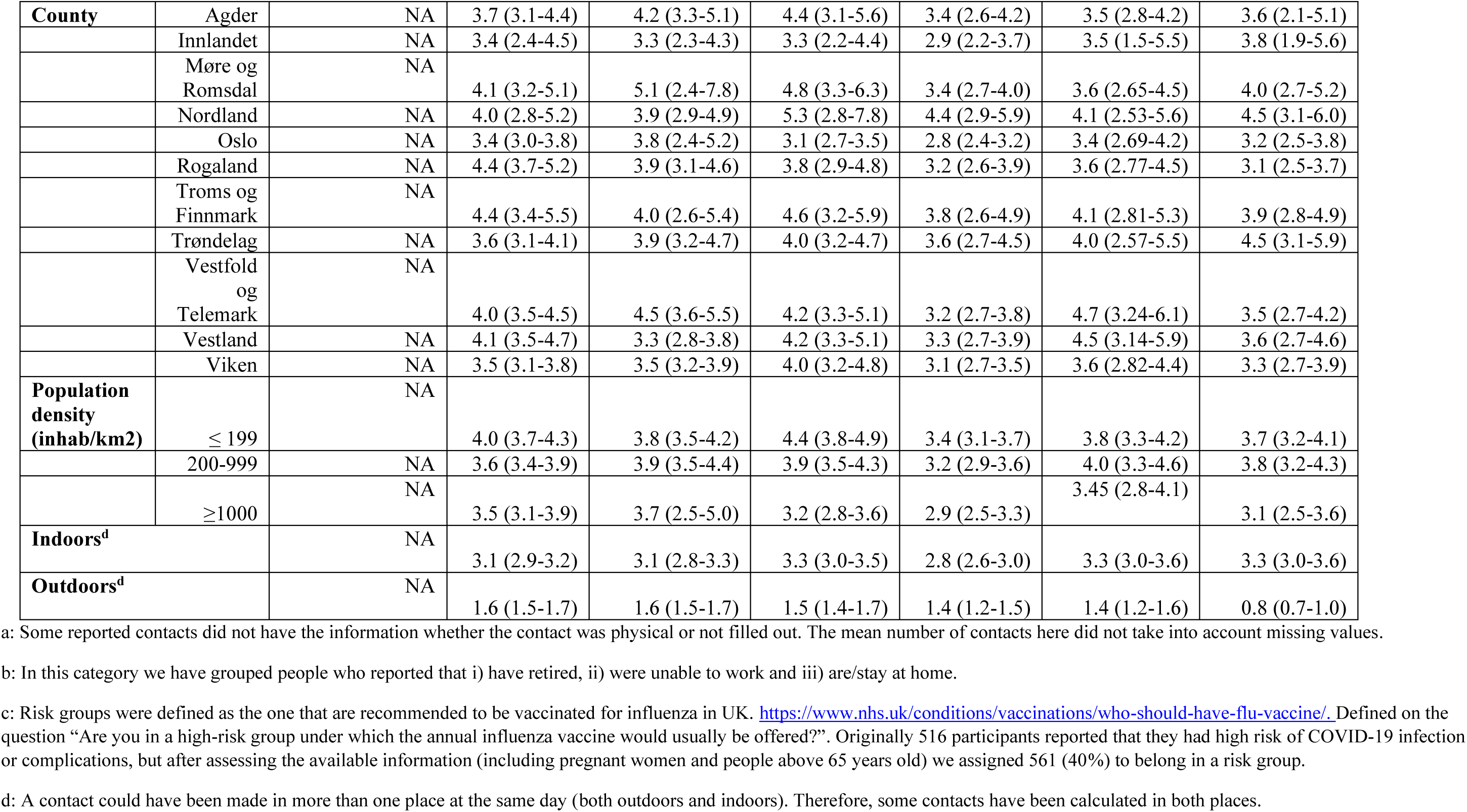
Summary of the mean number of daily contacts reported by adult participants (≥18 years) in the 2017 baseline survey and in each wave of the CoMix survey in 2020 (starting date of the data collection is indicated in brackets). These results are weighted by gender and age group.

**Fig. 2:**
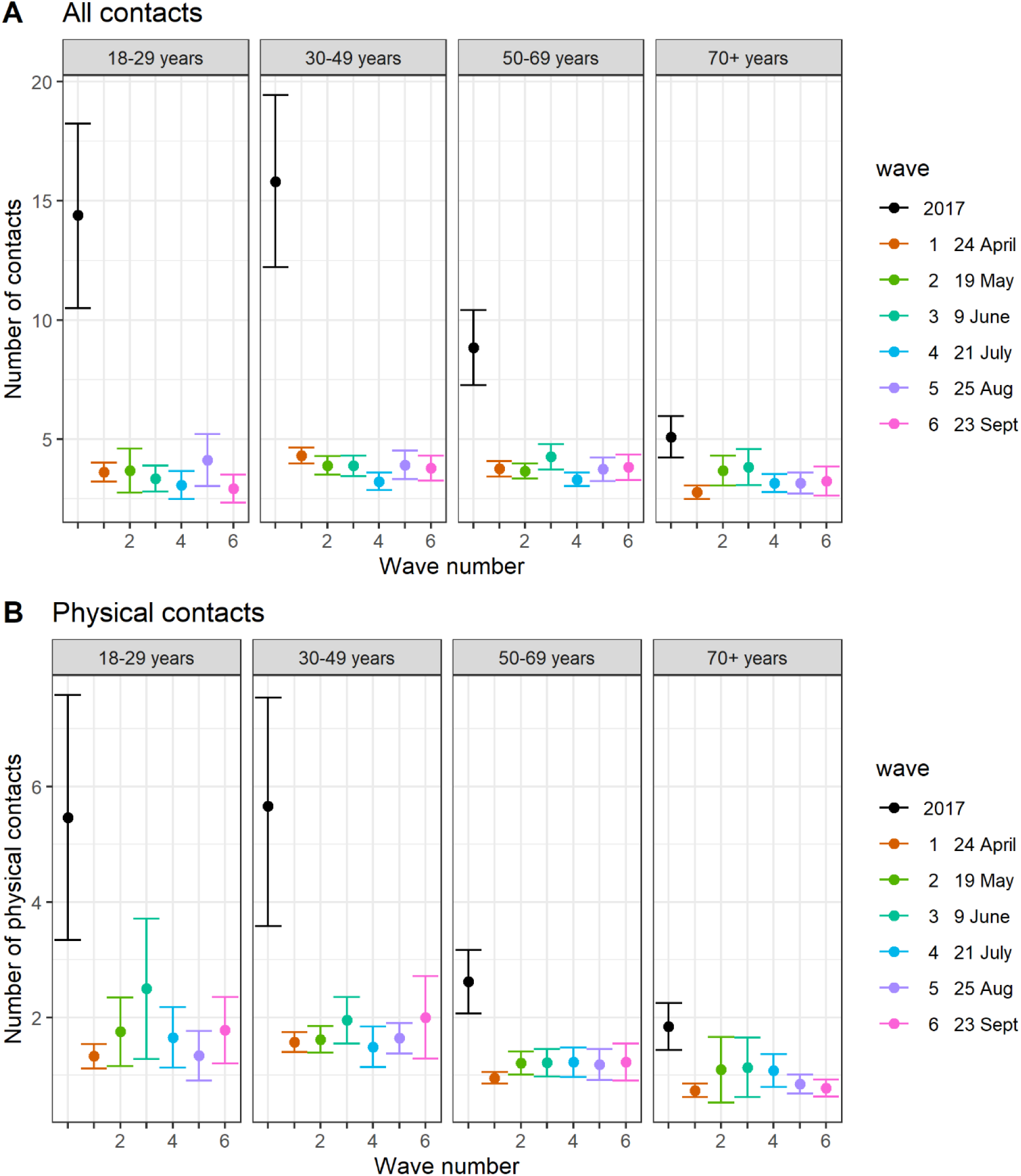
Mean number of daily contacts with 95% confidence interval stratified by participant age group for the 2017 baseline survey and the CoMix waves: (A) for all contacts and (B) for physical contacts reported. These results are weighted by gender.

The proportion of contacts involving touch (physical) declined during the early pandemic compared to pre-pandemic levels, fluctuating between 28 and 35% over the CoMix waves vs 38% in the baseline survey. For physical contacts, the mean number was lowest in April (1.2 per day, 95% CI 1.1-1.3), and highest in June (1.7 per day, 95% CI 1.4-2.0), corresponding to a drop of 73% and 62%, respectively. The age-specific changes were most prominent in younger adults, similar to that of all contacts (including physical and non-physical) (Figure 2B).

Participants reported significantly fewer contacts during the summer holiday. Aside from this observation, there were few significant differences between the waves. This is noteworthy, considering that social distance measures were relaxed during the late April-June period (see Online Resource, Section 3 for the timeline of control measures) and the low incidence of COVID-19 infections during the CoMix survey (Figure 1B). However, contacts among students (daily activity: school/education) that were low before the summer due to the closure of educational institutions decreased from August to September as digital teaching was changed back to in person teaching.

Persistent differences in contacts between weekdays and weekends were not observed. However, panellists reported fewer contacts during weekdays than weekends (Saturday-Sunday) in July-August (Table 2). Contacts made at home primarily reflected the household size, and the relative reduction of contacts in the summer holiday period (wave 4) was most pronounced for the smallest households. Indoor contacts constituted a significant proportion, accounting for 65% of all contacts reported in April and rising to 79% by the end of September (Online Resource, Fig. S1; Table S4). In terms of participant characteristics, employed participants reported significantly more contacts than participants with “at home” status across all Comix waves. The results were more mixed for participants under education (Table 2). There was no significant association found between participant gender, Norwegian-born status or geographic location and the reported number of contacts (Table 2).

### Changes in mixing pattern

The imputed CoMix contact matrices exhibit a general decline in contact rates when compared to the baseline. At the same time the mixing patterns observed in the baseline survey were maintained, characterised by high density along the diagonal (indicating mixing within the same age groups) and high-density “wings” (off-diagonals, representing mixing between children and their parents) (Figure 3). The location-specific matrices reveal a large degree of consistency across the waves. An exception is school contact rates, which remained consistently low, except for wave 5 in late August (Figure 4).

**Fig 3:**
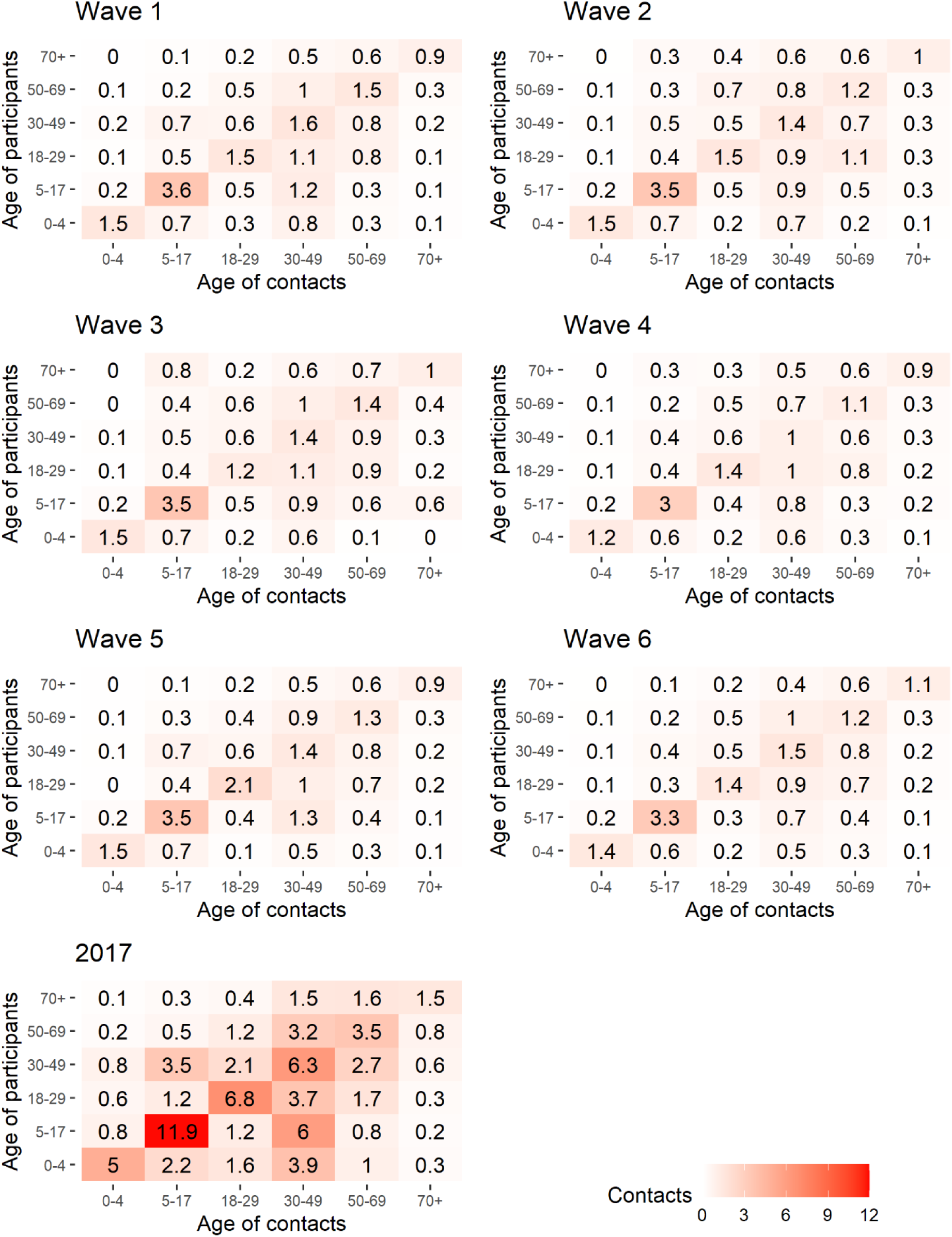
Imputed social contact matrices showing the mean daily number of contacts in the six CoMix waves. For comparison, the corresponding matrix from the 2017 baseline survey is shown below. The matrices represent bootstrap mean values of N=10 000 samples. Data were weighted on age and adjusted for reciprocity of contacts.

**Fig 4:**
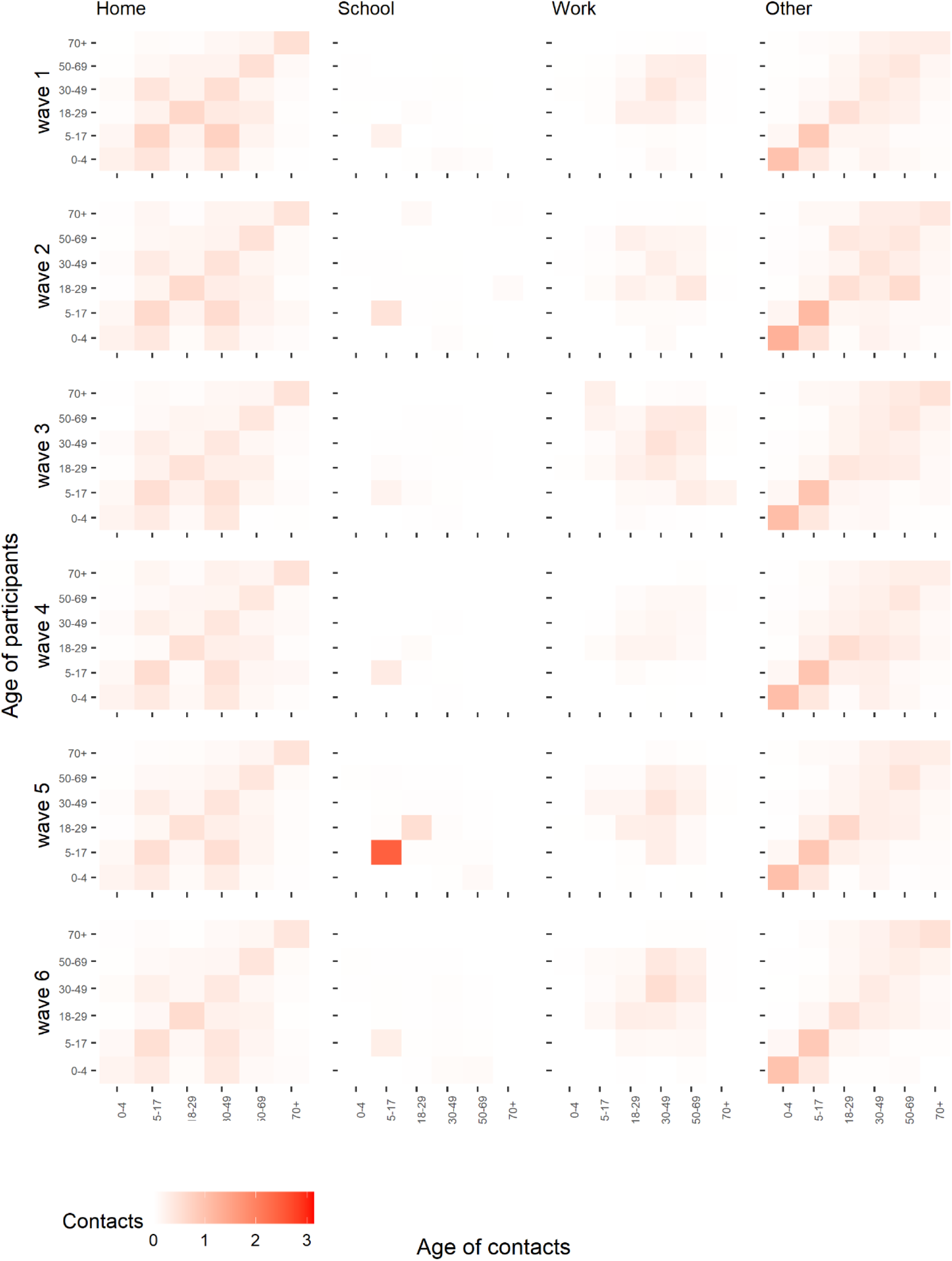
Imputed setting-specific social contact matrices showing mean number of daily contacts for the six waves in the CoMix survey. Locations include all contacts made in the home, at schools, at workplaces and other community contacts (transport, sport activities etc.). Note that some contacts overlap as multiple settings could be registered for the same contact. The matrices represent bootstrap mean values of N=10 000 samples. Data were weighted on age and adjusted for reciprocity of contacts.

Home contacts show mixing primarily with partners of the same age and children, and “other” community contacts display assortative mixing, particularly among the youngest age groups. The dominant eigenvectors of the CoMix matrices, which represents the contribution of different age groups to overall transmission, reveal a slightly more age-homogeneous contribution compared to the pre-pandemic matrix. However, in both surveys, school-age children made the most significant contribution when considering all contacts (see Figure 5A), and for physical contacts also the youngest children contribute significantly. (Figure 5B). The mean effective daily contact number (maximum eigenvalue) ranged from 3.7 to 4.6 in the CoMix waves, compared to 16.3 in the baseline survey (Figure 5C).

**Fig 5:**
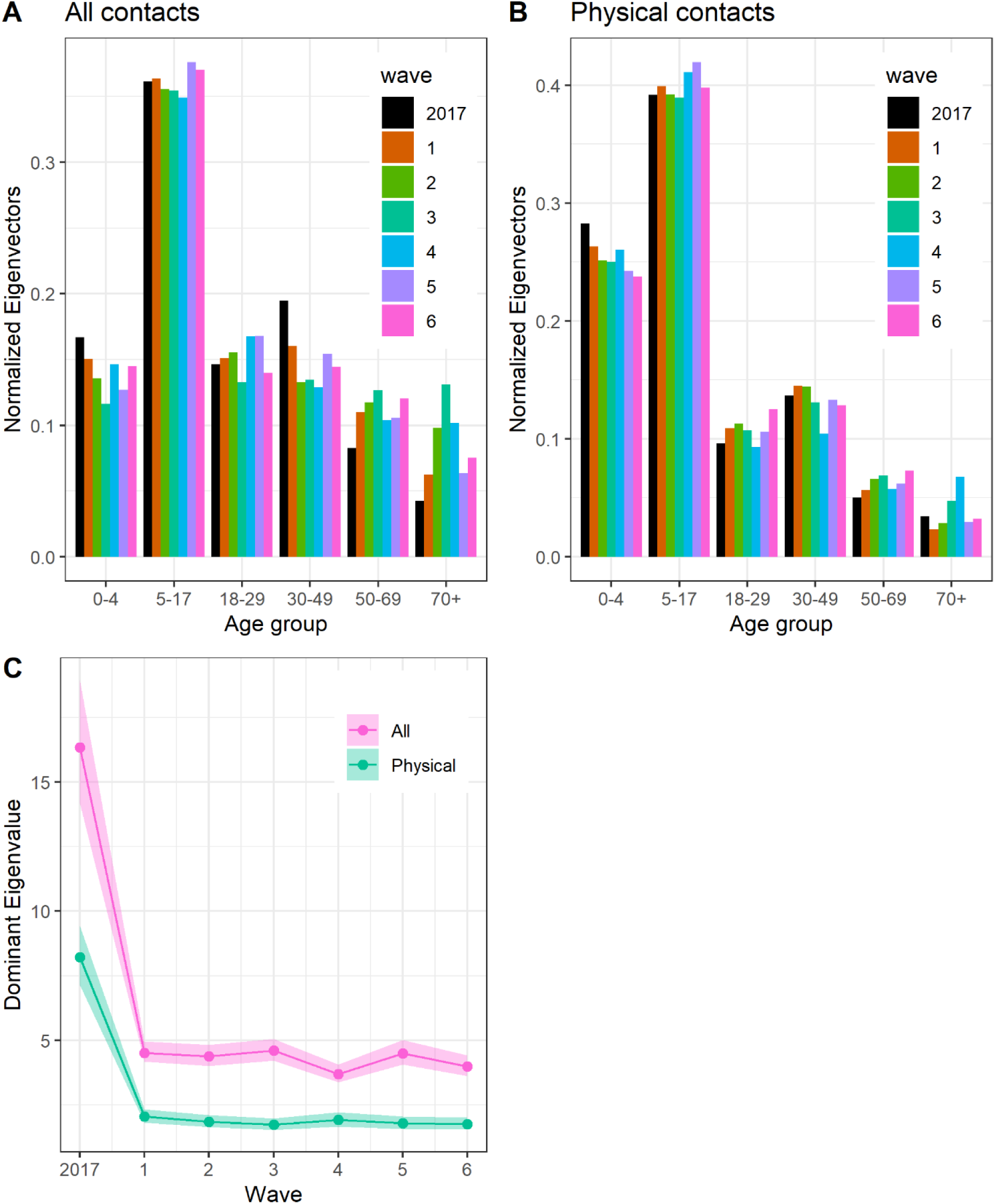
Comparison of CoMix and baseline dominant eigenvectors and eigenvalues: (A) Dominant normalised eigenvectors from contact matrices of all contacts by age group, (B) Dominant normalised eigenvectors from contact matrices of physical contacts by age group, (C) dominant (maximum) eigenvalues of contact matrices for all contacts (red) and physical contacts (green).

The index of disassortativity for all contacts was higher in the CoMix contact matrices, varying between 0.46 and 0.59, with the largest value in July, compared to 0.43 in the baseline survey. This suggests assortative mixing and a slight shift towards more intergenerational mixing during the early stages of the pandemic (Online Resource, Table S5). Regarding physical contacts, the disassortativity index varied from 0.46 to 0.54, compared to 0.5 in the baseline, indicating minimal change. By location, the most significant change was the notably higher values of the disassortativity index of school contacts in the CoMix study.

Overall, the CoMix contact matrices of physical contacts (Online Resource, Figure S5) were rather stable. These contacts were dominated by contacts in the home and ‘other’ settings (Online Resource, Figure S8), while contacts at school and workplaces were almost absent. The strong influence of household contacts is visible in the dominant eigenvectors where children (0 – 18 years) and their parents (30-50 years) were primary contributors to potential transmission, aligning with the baseline study (Figure 5B). The mean effective daily physical contact number varied between 1.7 and 2 throughout the survey, compared to 8.2 in the baseline (Figure 5C).

### Community contacts

Overall, the maximum eigenvalue ratios of the full population contact matrices categorised by type of contact and location showed a substantial reduction (Online Resource, Figure S4). The mean ratios of school contacts were consistently below 0.1, except in August when it reached 0.4 when considering all contacts. Work contact ratios displayed skewed distributions, but with mean levels typically around 0.2-0.3, except in July (wave 4), where they dropped to 0.1. The largest share of community contacts was of type “other”. Within this category, physical contacts peaked in July (wave 4), elevated by encounters associated with vacation activities, particularly contacts in other households, restaurants and bars, and other outside locations (Online Resource, Table S5). Contacts involving touch at school and workplaces remained low despite increasing contact numbers in August-September, indicating awareness and adherence to distancing recommendations.

### Estimated reproduction number

Figure 6 illustrates the estimated reproduction numbers during the survey period using the CoMix contact matrices for all contacts and physical contacts (essentially an estimate of the basic reproduction number R0). These estimates are presented alongside estimated values of the effective reproduction number from a calibrated model using Norwegian daily admission and test data (10, 22). The reproduction numbers based on overall contact matrices exhibited more variability across the waves compared to the ones based on physical contact matrices and showed a somewhat closer alignment with the model-derived estimates, including a dip in July and increasing transmission in August. However, both methods (based on overall or physical contact matrices) suggest potential overestimation in April and underestimation in September when compared to the real-time model-derived estimates.

**Fig 6:**
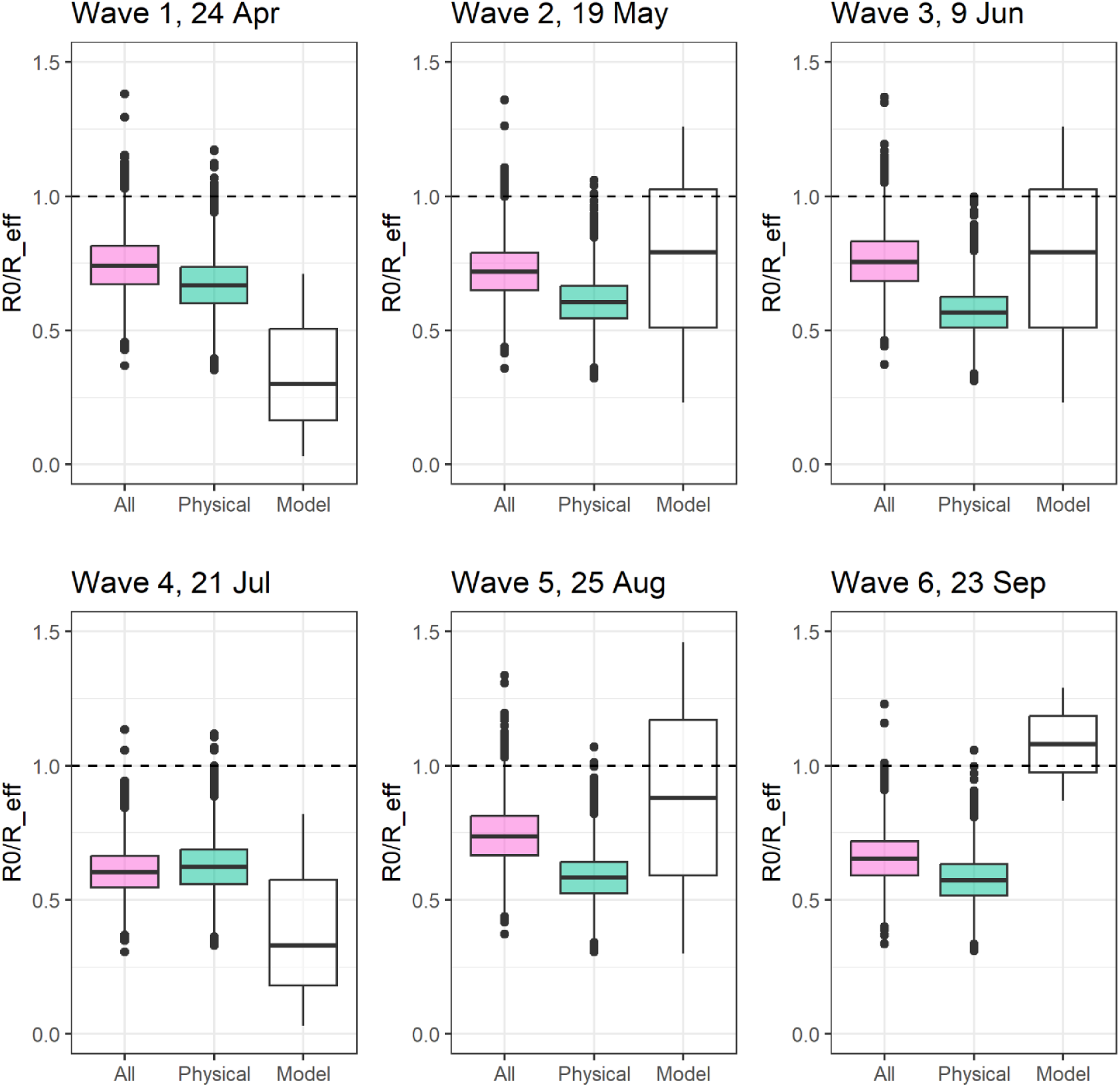
Boxplot showing estimated R0 reproduction numbers for the CoMix waves (all contact matrices and physical contact matrices), multiplying maximum eigenvalue ratios (Imp CoMix/Baseline) from imputed contact matrices with R0 from a calibrated mathematical model prior to the lockdown in Norway (10, 22). For comparison is shown the estimated effective reproduction numbers R_e (t) from the same model during the waves

### Personal protective behaviour

Use of personal protective measures, including hand washing, use of hand sanitiser or face masks can impact transmissibility. Hand hygiene in Norway has been strongly recommended since the start of the pandemic (see Online Resource, Table S3). In our survey, 95% of the participants reported that they had at least once washed their hands or used hand sanitiser in the last three hours, and this was a consistent finding over all six waves (range 94-96%). Only 3-4% of the participants in April-July reported using face masks, and this number increased to 6% and 9% in August and September, respectively. As a preventive measure for COVID-19, face masks were recommended from mid-August 2020 in the capital Oslo and the surrounding Østfold region during public transportation where a minimum of one-meter distance could not be met (5).

## Discussion

This study provides insights into the changes in social behaviour during the early COVID-19 pandemic in Norway. The CoMix survey started in April 2020, just two weeks after the end of the national lockdown that involved strict social distance measures, including the closure of schools, kindergartens, fitness centres, restaurants, hair salons, etc. While the measures gradually eased towards the summer, certain restrictions remained in place until the study finished in late September (4, 5).

We found a significant reduction of 67-73% in the mean number of daily contacts from April to September 2020 compared to a pre-pandemic baseline survey. This reduction was most pronounced among younger adults. The primary factor contributing to the decline was a decrease in community contacts, highlighting households as a major venue for ongoing transmission. Our findings are comparable to reductions in contacts reported from other countries in that period. In UK, a 79% decrease of contacts was reported during lockdowns, lowering to 57% in the summer when measures eased (23). In China, where interventions were stricter, 86% and 88% reductions were observed in February in Wuhan and Shanghai, respectively (24). In other countries with data until the autumn of 2020, the mean number of contacts increased significantly during the period (only small increase observed in Norway) following the lockdown or easing of social distance measures, including Belgium (69-80%), the Netherlands (41-72%), Luxemburg (59-82%), and the USA (60-82%) (20, 25–27); but still remained to levels below the pre-pandemic.

Interestingly, the Norwegian results showed no noticeable changes in social mixing patterns in response to the scale-back of restrictions, though, certain mitigation measures were maintained. This finding may be attributed to the high level of awareness in the Norwegian population and a general trust in authorities (28). During this period, the Ministry of Health together with the leadership of the Norwegian Institute of Public Health and the Norwegian Directorate of Health held televised press briefings one to three times per week to communicate restrictions and provide information about preventive measures (4). The low proportion of physical contacts at schools and workplaces throughout the survey suggests that employers and institutions effectively implemented measures. We estimated the effective mean number of daily contacts per person for the whole population to be around 4-5, and 2-3 for physical contact, corresponding to a drop of 69-78% in expected transmissibility compared to regular social interactions. In the baseline survey, mixing patterns during the early pandemic displayed assortative tendencies that changed somewhat onwards more inter-age group contacts. This shift was driven by a large drop in school contacts, work-related and leisure activity contacts, while contacts within households, which were more age-disassortative, declined less. Connections in the community were primarily in other premises, and these random contacts could potentially support connectivity between population groups (“small-world effects”).

Regarding the estimation of reproduction numbers, our results were broadly consistent with those of an independent model calibrated to Norwegian data (10). We found that using overall contact matrices as opposed to physical contact matrices gave higher level of consistency with the epidemiological dynamics. Furthermore, the physical contact matrices suggest an initial epidemic driven by children, akin to influenza, while a Norwegian serological survey from January 2021 found no differences in seroprevalence by age group (29). The Norwegian Comix study was used to gauge contact contributions from different settings in relation to parameterisation of individual-based models used in scenario analyses and planning purposes during the response to the pandemic.

Several other countries also started collecting contact pattern data early in the pandemic including the CoMix partners. The SOCRATES platform brings together the CoMix social contact data from over 20 European countries collected at different points in time throughout the COVID-19 pandemic (30). A comparison of the contact pattern data among countries has been published elsewhere highlighting the importance of these data in evaluating a diverse range of physical distancing control measures (15).

### Limitations

The low response rate in the online CoMix survey may have resulted in selection bias concerning behavioural characteristics, computer literacy, adherence to mitigation measures, and more. For example, Ipsos obtained only a 7% recruitment rate in wave 1. Despite anonymity, participants might have felt pressure to report fewer contacts due to the recommendations and restrictions that were in place, which was likely less the case during the baseline survey that took place during “peace time”. Additionally, systematic differences in sample populations and answers between the CoMix and baseline surveys may result from differences in design, recruitment methods, and type of media used for the questionnaire. While the gender balance was well-maintained in both surveys, there were clear differences in age profiles. For example, the baseline survey was more biased towards elderly, with 43% compared being older than 70 years compared to 16% in the CoMix survey. We believe the CoMix survey is fairly representative of a Norwegian adult population, but it may not be optimal for all aspects of comparative analyses with the baseline survey. We should note that the baseline survey had as well a low response rate of 12% also indicating the difficulties of conducting such studies.

Another limitation is the exclusion of children below 18 years in the CoMix survey to expedite the launch and avoid a prolonged process for ethical approval. Children constitute a significant portion of the population with distinct social behaviours. In this study, we addressed the absence of data on children’s contacts by imputation, re-scaling portion of contact matrices from the baseline study based on behavioural changes in adults. This procedure is potentially problematic because it assumes that children and adults respond similarly. Additionally, some control measures during the time of the study were designed to protect children’s usual activities more than adults and were therefore less strict. Although we have gauged children-to-adult contacts using the available information on adult-to-children contacts, it does not remedy the lack of data on children-to-children contacts.

Our reproduction number estimates did not consider depletion of susceptible, age-specific infectivity or susceptibility, or seasonal forcing. It critically assumes that social contact is an adequate surrogate measure of potential transmission events and that the contacts are comparable across the waves. However, other factors could have affected the risk of transmission during contact, such as personal hygiene measures (e.g., hand washing, face masks), environmental factors (e.g., ventilation, open areas), physical distance with conversational contacts, and the duration of contacts. For example, the potential overestimation of the reproduction number in April, measured against the modelling result, may be related to the early scare and heightened risk perception, which could have affected behaviour during contact. Conversely, the potential underestimation in September may be associated with increasing time spent indoors. However, the information on personal protective behaviour in our study suggested little change between waves.

Other findings from the Norwegian CoMix survey have underscored the importance of cognitive and psychological factors in predicting behaviour, in particular identifying predictors of visiting or intending to visit crowded places during the study period (18). Furthermore, the information gathered through social surveys can be complemented by research utilising non-standard data sources. These may include data streams such as mobile phone tracking to monitor movement patterns (31). Such multi-faceted research is essential for enhancing our understanding of the dynamic interaction between physical distancing measures, disease prevalence, and population response.

Despite the limitations, our results contribute valuable empirical information into the social behaviour of the Norwegian population during the early COVID-19 pandemic when significant social distancing measures were in place. Social contact data is critical for public health authorities to monitor behaviour, adherence, and have played a pivotal role in developing models used to guide COVID-19 policies. This information will also prove useful in future crisis situations when similar mitigation measures may be required.

## Data Availability

The dataset analysed for the study contained anonymized individual-level data. All data were stored securely, and confidentiality was protected in accordance with the Data Protection Act, GDPR and in accordance with requirements of the Norwegian Health Research Act. The anonymised CoMix data used for the analyses will be available on the CoMix platform soon after the article is published.

## Acknowledgements

First and foremost, the authors wish to thank all the participants to the surveys in 2020 and 2017. The authors also acknowledge the efforts of Kiesha Prem from London School of Hygiene and Tropical Medicine and Niel Hens of the university of Antwerp for their work in preparing the European-wide protocols for the CoMix study, and Kaya Sinem Cetin who contributed to the first translation of the CoMix questionnaire into Norwegian. We also thank Sara Grant-Vest, Eva Voukelatou, Kate Duxbury and the Ipsos team that was involved in collecting the CoMix data. Regarding the survey in 2017, we acknowledge the contributions of our colleagues at the Department of IT Systems at Norwegian Institute of Public Health in Bergen (SIBIDI) that conducted the data collection.

## Statements & Declarations

### Ethics approval

The 2020 CoMix study (reference number 128391) and the 2017 study (REK 2016/385) in Norway were approved by the Regional Ethical Committee West. Individuals participated voluntarily in the survey and gave informed consent before inclusion. Data analyses were conducted on anonymised data.

### Consent to participate

Informed consent was obtained from all participants before inclusion in the study. Participants were informed in advance that the anonymised data would become publicly available at the end of the study. Participants had the right to request the withdrawal of their data at any time.

### Author contributions

LV, BR, AS and BFD in collaboration with members of the international CoMix team including CIJ, AG, WJE, KVZ planned the study, supervised data collections, and were responsible for data facilitation. BR and BFB were the project managers for the Norwegian CoMix study. LV and BFB coordinated the CoMix study, conducted the statistical analysis, and drafted the manuscript. LV, AS, BAW, DFV and BFD planned and coordinated the 2017 study. All co-authors contributed to the interpretation of the results. All co-authors contributed to the revision of the manuscript and approved the final version for submission. The corresponding author attests that all listed authors meet authorship criteria and that no others meeting the criteria have been omitted.

