## supplement material for "Social contact patterns during the early COVID-19 pandemic in Norway: insights from a panel study, April to September 2020"

### Article Title:

### 1. Group contacts

From wave 2 to 6, 453 participants reported group contacts (449 of those also reported individual contacts); ranging from 11% to 19% of the individuals with contact data per wave. We noticed that some participants reported high numbers with few of them reporting extreme values, such as in wave 2 where there were two participants that reported 4 847 and 7 021 contacts each. As mentioned in the main article the group contacts were excluded in the main analysis.

As an additional analysis to assess the impact of group contacts in wave 2 to 6, we calculated the mean number of contacts after fixing the maximum number of group contacts at 50, thereby reducing the influence of participants with exceptionally high contact numbers to the population mean. This affected 12 individuals in wave 216 in wave 3, 8 in wave 4, 16 in wave 5 and 12 in wave 6, all of whom had reported more than 50 group contacts. We present the results from this analysis in Table S1. When comparing these figures with the mean contact numbers reported in Table 2, we observed an increase of 1,2 to 3,1 on average throughout waves 2 to 6. Similar as in Table 2, the lowest number of contacts was reported in wave 4 and the highest in wave 3.

**Table S1:** Summary of daily individual and group contacts reported in each wave of the CoMix survey in 2020. The crude mean of contacts (not weighted) per wave is presented in the last column.

| CoMix Wave | Participants | Group contacts | Individual contacts | Total contacts | Crude mean of contacts |
|---|---|---|---|---|---|
| **Wave 1** | 1 400 | N.A.* | 5 360 | 5 360 | 3.8 |
| **Wave 2** | 1 182 | 2 586 | 4 477 | 7 063 | 6.0 |
| **Wave 3** | 1 012 | 3 020 | 4 041 | 7 061 | 7.0 |
| **Wave 4** | 931 | 1 144 | 3 011 | 4 155 | 4.5 |
| **Wave 5** | 768 | 1 839 | 2 850 | 4 689 | 6.1 |
| **Wave 6** | 645 | 1 678 | 2 335 | 4 013 | 6.2 |

**N.A.: Not available. In wave 1, participants were not able to report group contacts.*

***Total contacts= Sum of group and individual contacts.*

### 2. Representativeness of study population

Our participants were sampled to be representative by age, gender, and region of residence. The dropouts and new recruitment of participants impacted a little in the composition of the sample with only small changes in the representativeness (Table S2).

We observed that the representativeness by gender and county was quite good. Regarding the age distribution, the 18-30 age group were undersampled ranging from 7 to 16% across the waves compared to the 20% representation in the general population. Conversely, the 50-70 age group showed a slight oversampling, ranging from 35 to 40% through the waves in contrast to the 30% representation in the general population. In addition, participants residing in Oslo County were marginally overrepresented throughout the waves, constituting 17-18% of the sample compared to the 13% presentation in the general population. While these variances were considered minor for the purpose of our study, we decided to conduct weighted analysis for the contact pattern data to account for these differences.

**Table S2:** Sample characteristics in the baseline survey and of each wave of the CoMix survey (starting date of the data collection is indicated) compared to the proportions of the 2020 Norwegian adult population.

| **Demographic characteristics** | **Number of participants (%)** | | | | | | | **% of the adult Norwegian population 2020** |
|---|---|---|---|---|---|---|---|---|
| | **Baseline survey, 2017 n=309** | **Wave 1 (24 April), n=1400** | **Wave 2 (19 May), n=1182** | **Wave 3 (9 June), n=1012** | **Wave 4 (21 July), n=931** | **Wave 5 (25 Aug), n=768** | **Wave 6 (23 Sept), n=645** | |
| **Gender** | | | | | | | | |
| Male | 146 (47 %) | 702 (50 %) | 610 (52 %) | 534 (53 %) | 487 (52 %) | 421 (55 %) | 357 (55 %) | 50 % |
| Female | 163 (53 %) | 698 (50 %) | 569 (48 %) | 475 (47 %) | 441 (47 %) | 345 (45 %) | 287 (45 %) | 50 % |
| NA | 0 | 0 | 3(-) | 3(-) | 3(-) | 2 (-) | 12 (-) | - |
| **Age group** | | | | | | | | |
| 18-29 | 46 (15 %) | 217 (16 %) | 121 (10 %) | 98 (10 %) | 78 (8 %) | 57 (7 %) | 70 (11 %) | 20 % |
| 30-49 | 36 (12 %) | 509 (36 %) | 410 (35%) | 352 (35 %) | 300 (32 %) | 257 (34 %) | 210 (33 %) | 34 % |
| 50-69 | 93 (30 %) | 493 (35 %) | 458 (39%) | 406 (40 %) | 406 (44 %) | 319 (42 %) | 262 (41 %) | 30 % |
| 70+ | 134 (43 %) | 181 (13 %) | 193 (16 %) | 156 (15 %) | 147 (16 %) | 135 (18 %) | 103 (16 %) | 16 % |
| **County** | | | | | | | | |
| Agder | NA | 70 (5 %) | 57 (5 %) | 53 (5 %) | 42 (5 %) | 41 (5 %) | 29 (5 %) | 6 % |
| Innlandet | NA | 61 (4 %) | 51 (4 %) | 46 (5 %) | 54 (6 %) | 35 (5 %) | 28 (4 %) | 7 % |
| Møre og Romsdal | NA | 66 (5 %) | 58 (5 %) | 47 (5 %) | 49 (5 %) | 42 (6 %) | 38 (6 %) | 5 % |
| Nordland | NA | 50 (4 %) | 47 (4 %) | 43 (4 %) | 32 (3 %) | 24 (3 %) | 24 (4 %) | 5 % |
| Oslo | NA | 254 (18 %) | 209 (18 %) | 186 (18 %) | 156 (17 %) | 132 (17 %) | 113 (18 %) | 13 % |
| Rogaland | NA | 124 (9 %) | 105 (9 %) | 88 (9 %) | 82 (9 %) | 75 (10 %) | 66 (10 %) | 9 % |
| Troms og Finnmark | NA | 62 (4 %) | 44 (4 %) | 42 (4 %) | 36 (4 %) | 37 (5 %) | 21 (3 %) | 5 % |
| Trøndelag | NA | 131 (9 %) | 110 (9 %) | 90 (9 %) | 73 (8 %) | 58 (8 %) | 52 (8 %) | 9 % |
| Vestfold og Telemark | NA | 114 (8 %) | 94 (8 %) | 79 (8 %) | 81 (9 %) | 61 (8 %) | 53 (8 %) | 8 % |
| Vestland | NA | 167 (12 %) | 136 (12 %) | 113 (11 %) | 111 (12 %) | 82 (11 %) | 80 (12 %) | 12 % |
| Viken | NA | 301 (22 %) | 271 (23 %) | 225 (22 %) | 215 (23 %) | 181 (24 %) | 141 (22 %) | 23 |

### 3. Control measures overview during the study period

In Norway, the stringency of control measures implemented since the beginning of the pandemic varied depending on the evolving epidemiological situation. In Table 3, we present some of the social distance measures implemented by the Norwegian government during the data collection period of the Comix study. We should note that the social distance measures included in Table S3 are referring to the national guidelines, and there may have been variations in local measures may varied in some instances. Additionally, individual companies may have implemented their own distinct measures.

**Table S3:** Control measures implemented during the week of the CoMix data sampling (starting date of the data collection is indicated) and COVID-19 weekly incidence in Norway, April-September 2020.

| | **Data collection period** | | | | | | **Dates during Comix Study that the measures were implemented** |
|---|---|---|---|---|---|---|---|
| **Control measures** | **Wave 1, 24 April** | **Wave 2, 19 May** | **Wave 3, 9 June** | **Wave 4, 21 July** | **Wave 5, 25 Aug.** | **Wave 6, 23 Sept.** | |
| Ban all events | Yes | No | No | No | No | No | 12/3/2020-6/5/2020 |
| Closure of kindergartens/daycare | No | No | No | Summer break | No | No | 12/3/2020-20/4/2020 |
| Closure of primary schools- | Yes until 26 April | No | No | Summer break | No | No | 12/3/2020-26/4/2020 |
| Closure of higher education/universities | Yes | Yes | Yes until 14th June | Summer break | No | No | 12/3/2020-14/6/2020 |
| Closure of secondary schools | Yes | No | No | Summer break | No | No | 12/3/2020-11/5/2020 |
| Closure of cafes-restaurants | Yes | Yes | No, but with restrictions* | No, but with restrictions* | No, but with restrictions* | No, but with restrictions* | 12/3/2020-31/5/2020, Open but with restrictions* after 1/6/2020 |
| Closure of pubs/bars | Yes | Yes | No, but restrictions applied* | No, but with restrictions* | No, but with restrictions* | No, but with restrictions* | 12/3/2020-31/5/2020, Open but with restrictions* after 1/6/2020 |
| Closure of gyms, sports centres | Yes | No | No | No | No | No | 12/3/2020-14/5/2020 |
| Any gatherings above 50, indoors or outdoors not allowed | Yes | No | No | No | No | No | 7/5/2020-14/6/2020 |
| Teleworking strongly suggested | Yes | Yes | Yes | No | No | No | 10/3/2020-17/6/2021 |
| Private gathering restrictions | Yes (Advice to not meet in groups >5) | Yes (Advice to not meet in groups >20) | Yes (Advice to not meet in groups >20) | Yes (Advice to not meet in groups >20) | Yes (Advice to not meet in groups >20) | Yes (Advice to not meet in groups >20) | From 12/3 advised not to meet in groups of more than 5. Also advised to meet outdoors. From 7/5 advice to avoid gathering >20 people in private homes. |
| **Weekly Reported cases among all Norwegian population (sampling week)** | 359 (week 17) | 101 (week 21) | 80 (week 24) | 94 (week 30) | 374 (week 35) | 776 (week 39) | |

**Note: From 1[st] of June, cafes/restaurants/bars could reopen as long as they could provide a distance of one meter between guests and seating for all guests. All places had to close by 24.00.*

### 4. Social Contact patterns

#### 4.1 Additional data on number of contacts

**Fig. S1:** Mean number of reported daily contacts that occurred A) outdoors and B) indoors, stratified by participant age group and data collection period (wave). The starting date of the data collection is indicated in the brackets for data collected in the CoMix survey waves.

A)

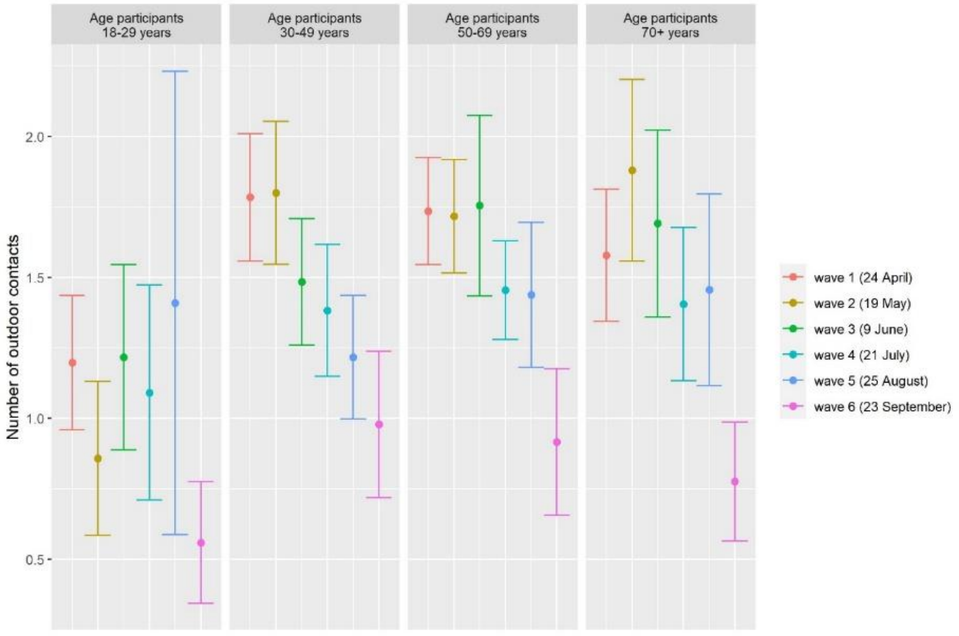

B)

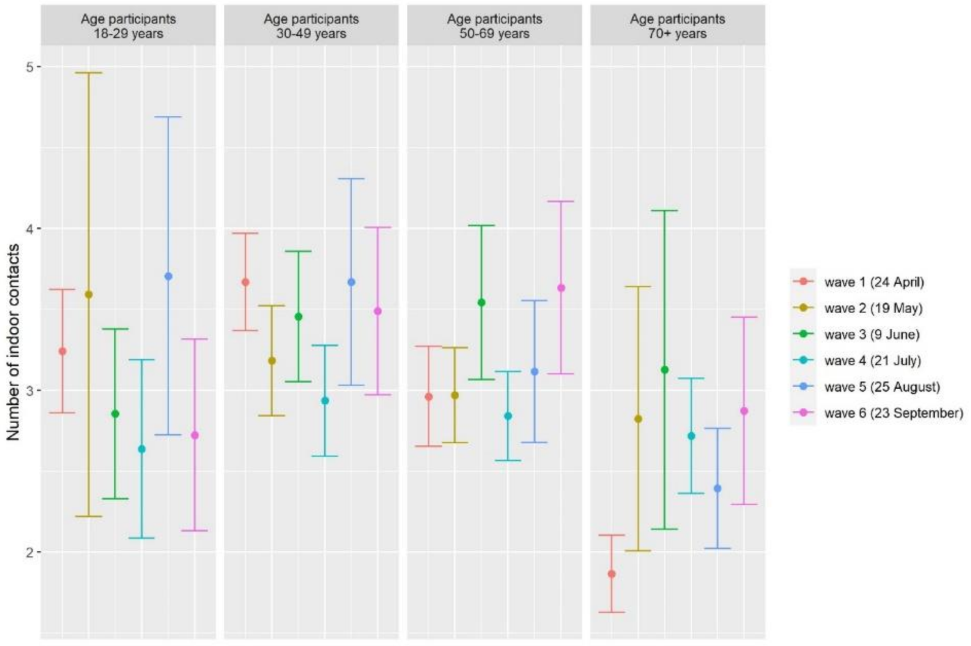

**Table S4:** Percentage (crude) of contacts that all participants reported by place* and type of contact in each wave.

| Place of contact and type of contact | | *Wave 1, 24–30 April, n=1400* | | *Wave 2, 19–26 May, n=1182* | | *Wave 3, 9–16 June, n=1012* | | **Wave 4, 21–27 July, n=931** | | **Wave 5, 25 Aug–2 Sept,` n=768** | | **Wave 6, 23–30 Sept, n=645** | |
|---|---|---|---|---|---|---|---|---|---|---|---|---|---|
| | | Total contacts | % | Total contacts | % | Total contacts | % | Total contacts | % | Total contacts | % | Total contacts | % |
| Place of contact | Home | 2 457 | **41%** | 1 906 | **37%** | 1 401 | **31%** | 1 369 | **38%** | 1 075 | **33%** | 821 | **31%** |
| | Other house | 471 | **8%** | 566 | **11%** | 318 | **7%** | 397 | **11%** | 256 | **8%** | 230 | **9%** |
| | Work | 1 206 | **20%** | 784 | **15%** | 1 284 | **28%** | 390 | **11%** | 669 | **21%** | 714 | **27%** |
| | Supermarket-shops | 508 | **8%** | 682 | **13%** | 525 | **12%** | 499 | **14%** | 360 | **11%** | 262 | **10%** |
| | Outside other (parks-countryside) | 642 | **11%** | 490 | **10%** | 339 | **7%** | 270 | **8%** | 232 | **7%** | 115 | **4%** |
| | Public_transport | 104 | **2%** | 137 | **3%** | 116 | **3%** | 143 | **4%** | 86 | **3%** | 77 | **3%** |
| | School | 90 | **1%** | 104 | **2%** | 52 | **1%** | 18 | **1%** | 120 | **4%** | 64 | **2%** |
| | Leisure (bars, restaraunts etc) | 31 | **1%** | 131 | **3%** | 118 | **3%** | 157 | **4%** | 85 | **3%** | 85 | **3%** |
| | Sport | 66 | **1%** | 32 | **1%** | 42 | **1%** | 33 | **1%** | 107 | **3%** | 66 | **3%** |
| | Other | 466 | **8%** | 323 | **6%** | 357 | **8%** | 318 | **9%** | 266 | **8%** | 204 | **8%** |
| | **Total, all places[a]** | **6 041** | **100%** | **5 155** | **100%** | **4 552** | **100%** | **3 594** | **100%** | **3 256** | **100%** | **2 638** | **100%** |
| Physical contact | | 1 707 | **32%** | 1 307 | **29%** | 1 055 | **26%** | 1 062 | **35%** | 811 | **28%** | 672 | **29%** |
| Contact indoors[b] | | 4 389 | **65%** | 3 630 | **64%** | 3 407 | **68%** | 2 635 | **67%** | 2 452 | **70%** | 2 172 | **79%** |
| Contact outdoors | | 2 325 | **34%** | 2 007 | **36%** | 1 611 | **32%** | 1 296 | **33%** | 1 042 | **30%** | 576 | **21%** |

*a: A contact could have been made in more than one place at the same day. Therefore, the proportions were calculated using the total of contacts for all places reported.*

*b: A contact could have been made in more than one place at the same day (outdoors and indoors). Therefore, the proportions were calculated using the total of contact for both places.*

**Fig. S2:** Percentage (crude) of daily contacts reported by location* of participants in each wave. The starting date of the data collection is indicated in the brackets for data collected in the CoMix survey waves.

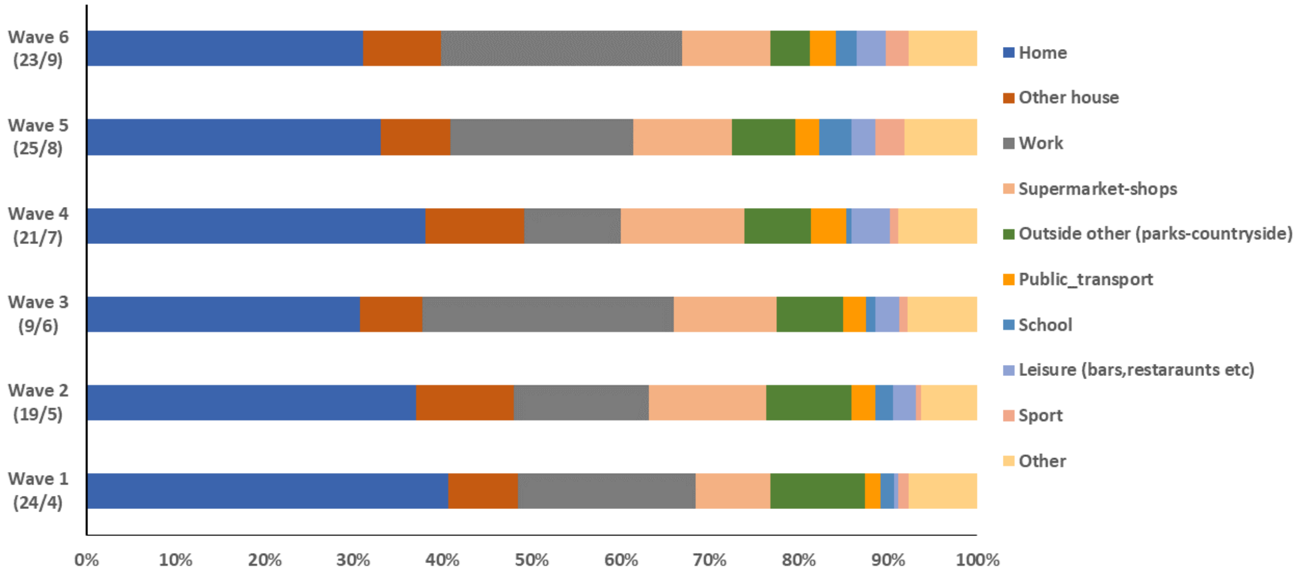

**Note: A contact could have been made in more than one place at the same day. Therefore, the proportions here were calculated using the total of contacts for all places reported.*

### 4.2 Contact matrices

**Fig S3:** Boxplots of the estimated ratio of the maximum eigenvalues (CoMix/Baseline) of the adult-to-adult contract matrices for each wave by location and type of contact. The ratios of dominant eigenvalues represent bootstrap sample pairs (N=10 000) that were used in the imputation process to scale contacts of children in the baseline contact matrices.

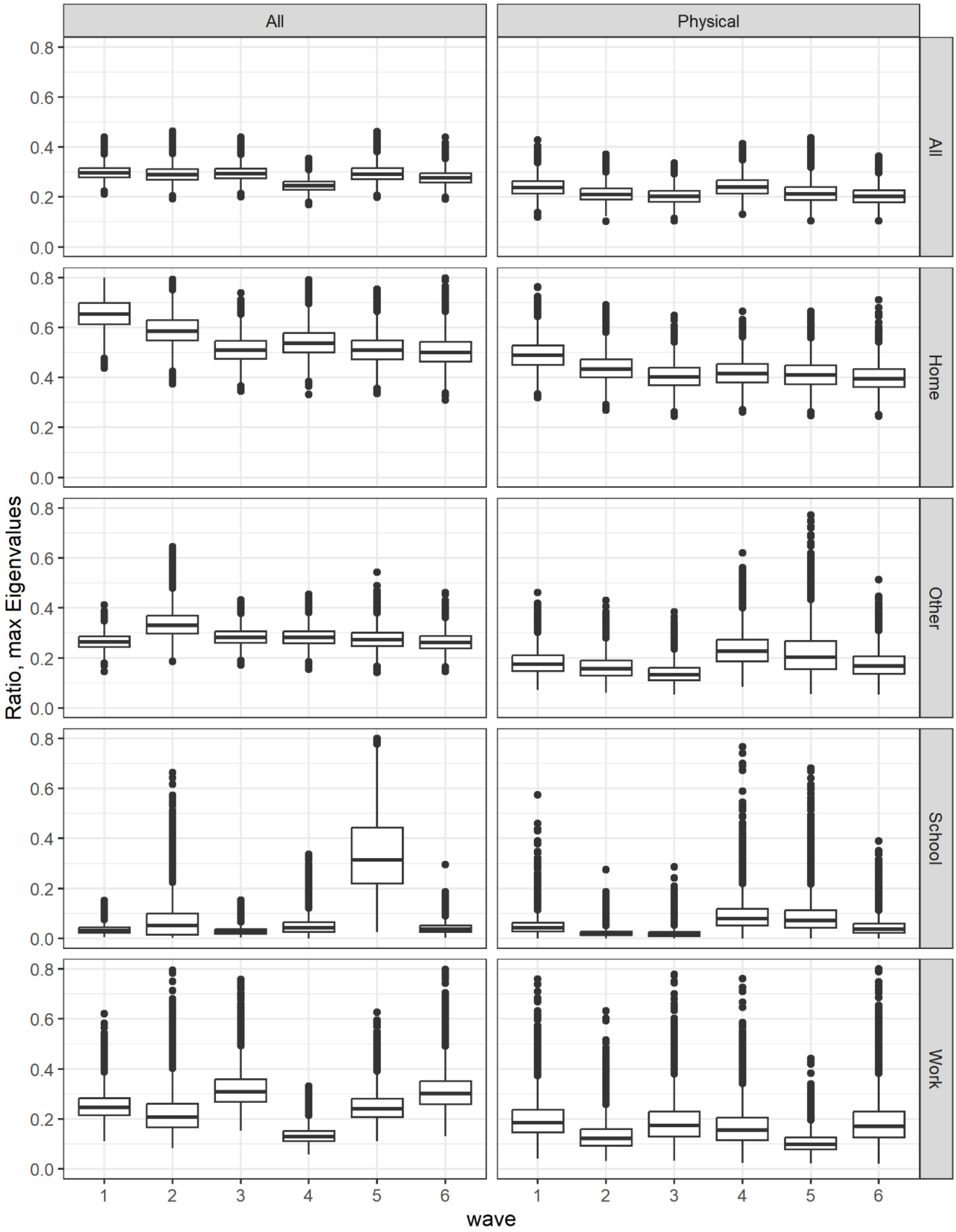

**Fig S4:** Boxplots of the estimated ratio of the maximum eigenvalues (CoMix/Baseline) of the full population matrices for each wave by location and type of contact. The ratios of dominant eigenvalues represent bootstrap sample pairs (N=10 000).

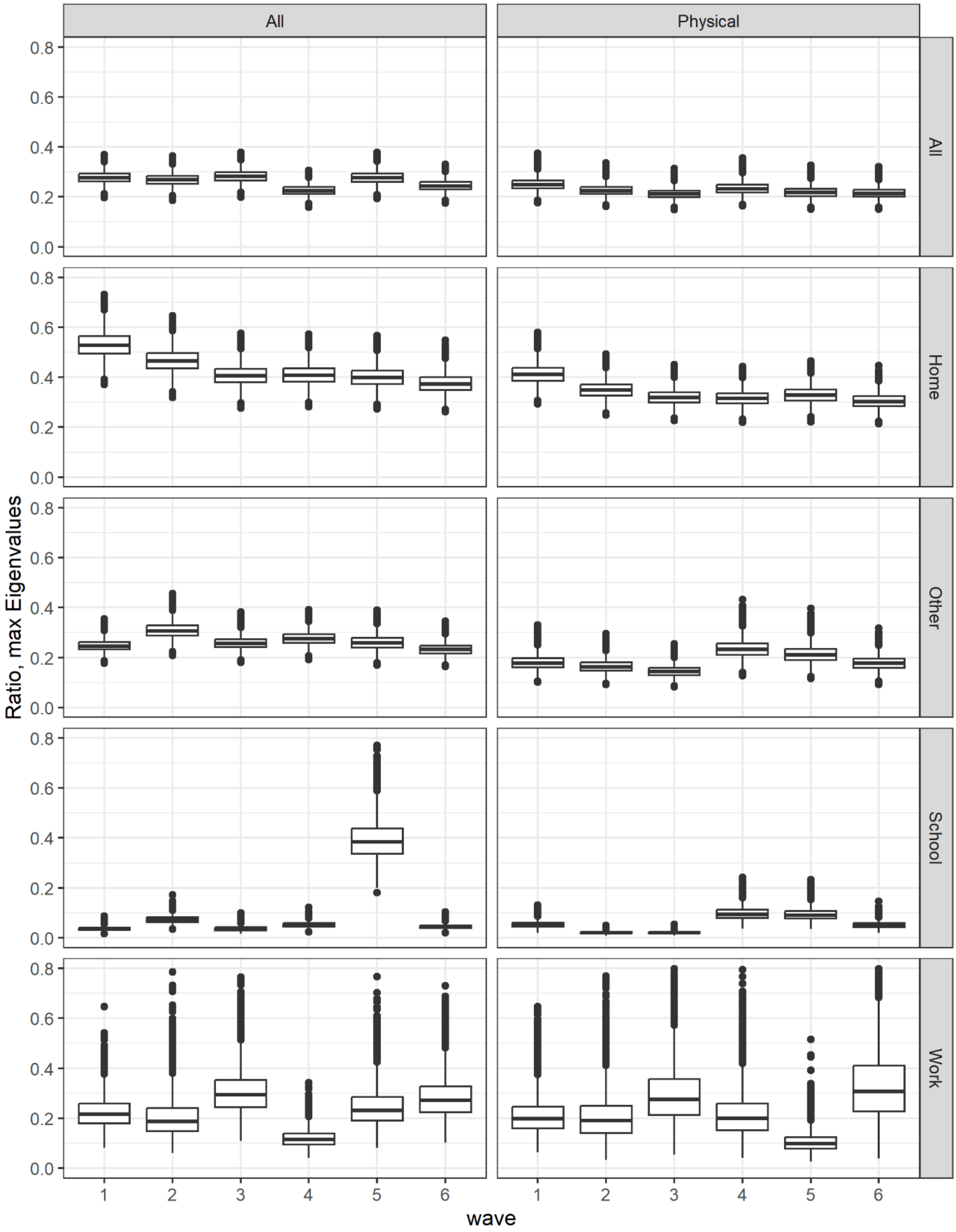

**Fig S5:** Imputed social contact matrices showing the mean number of daily physical contacts in the six CoMix waves; the corresponding matrix from the 2017 baseline survey is shown below as a reference. The matrices report bootstrap mean values from N=10000 samples. Data were weighted on age and adjusted for reciprocity of contacts.

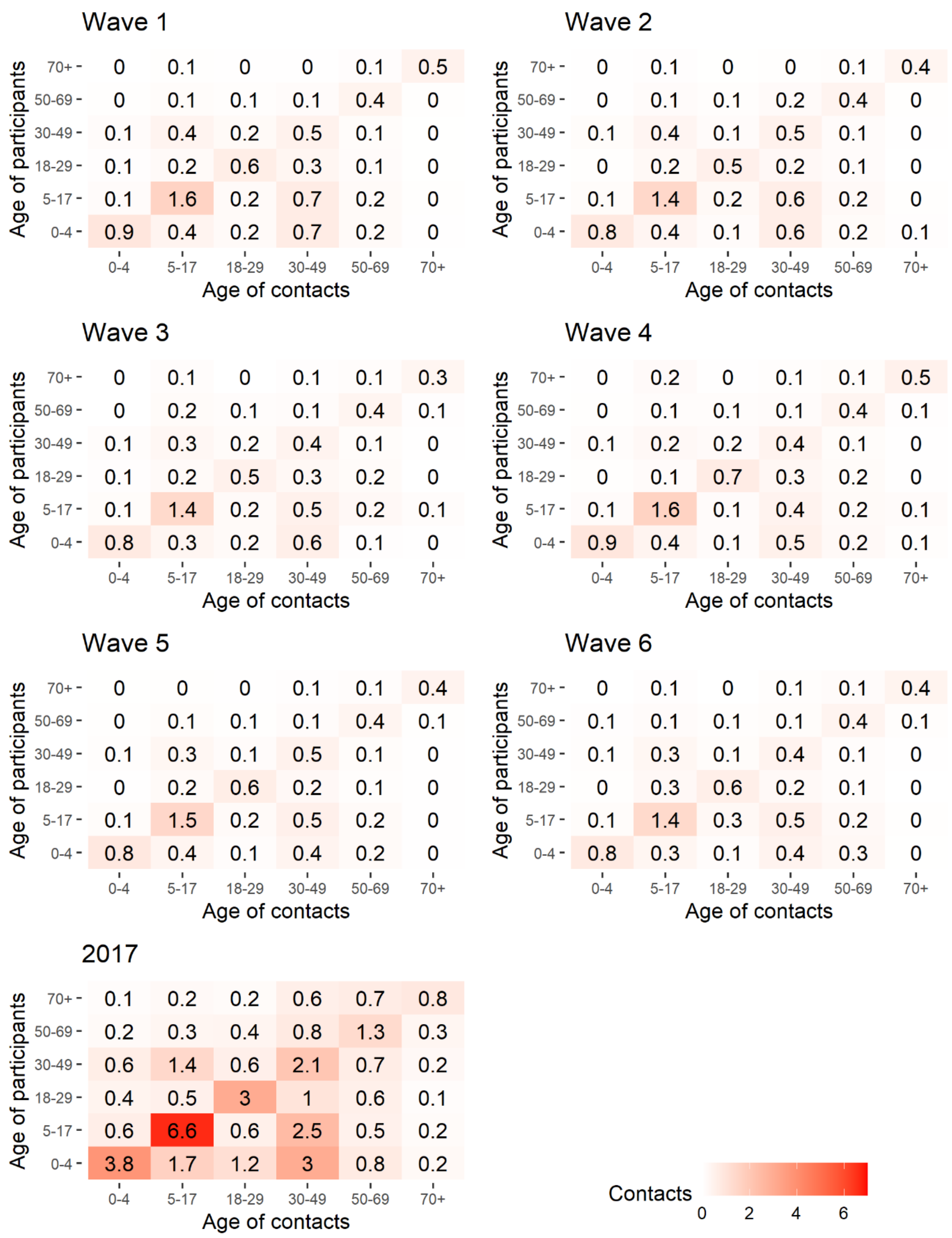

**Fig S6:** Social contact matrices of the six CoMix waves showing the mean number of daily contacts reported; below is shown the corresponding matrix of contacts reported by adults in the 2017 baseline survey as a reference. The figures represent bootstrap mean values from N=10 000 samples. Data were weighted on age.

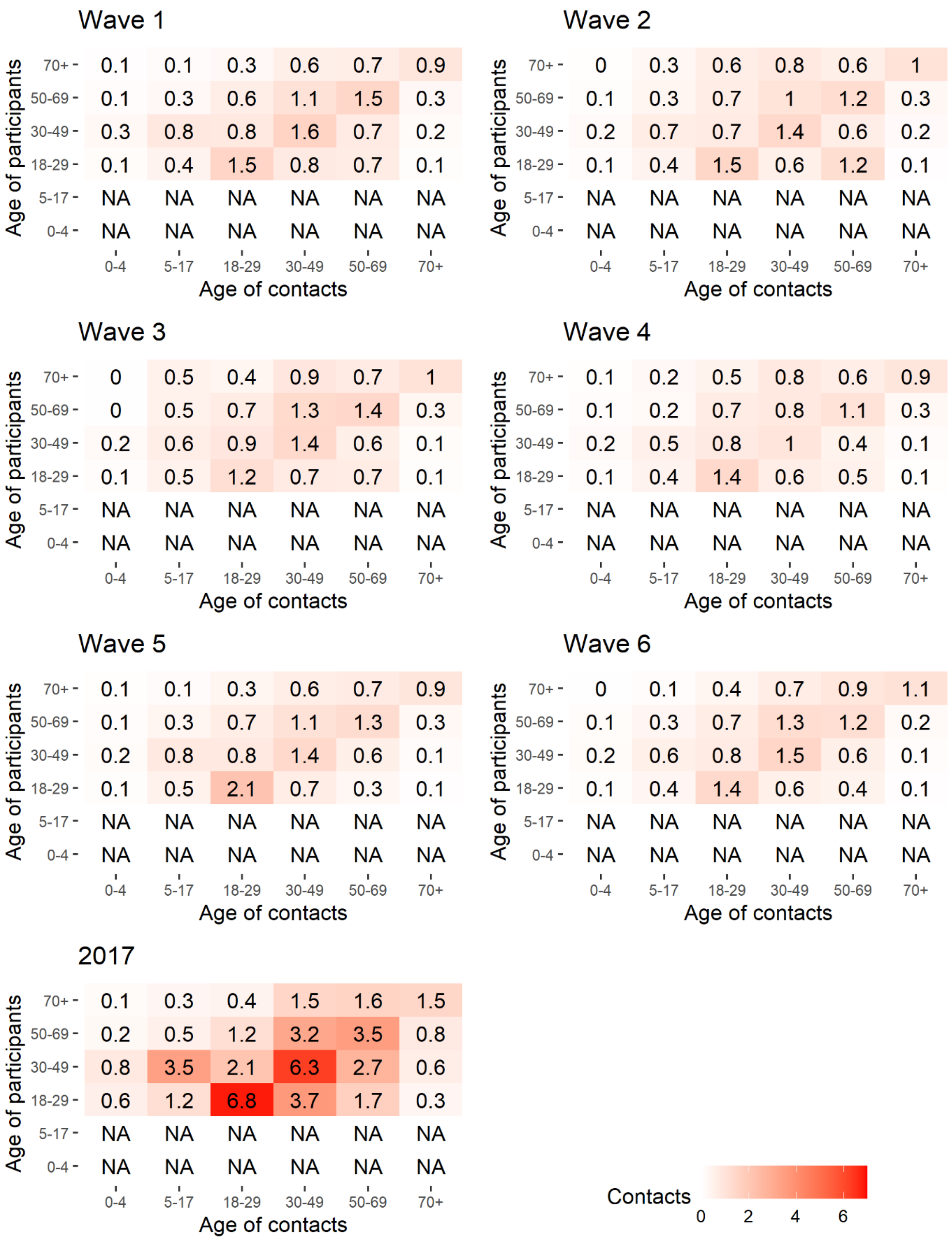

**Fig S7:** Social contact matrices of the six CoMix waves showing the mean number of daily physical contacts reported; below is shown the corresponding matrix of physical contacts reported by adults in the 2017 baseline survey as a reference. The matrices represent bootstrap mean values from N=10 000 samples. Data were weighted on age.

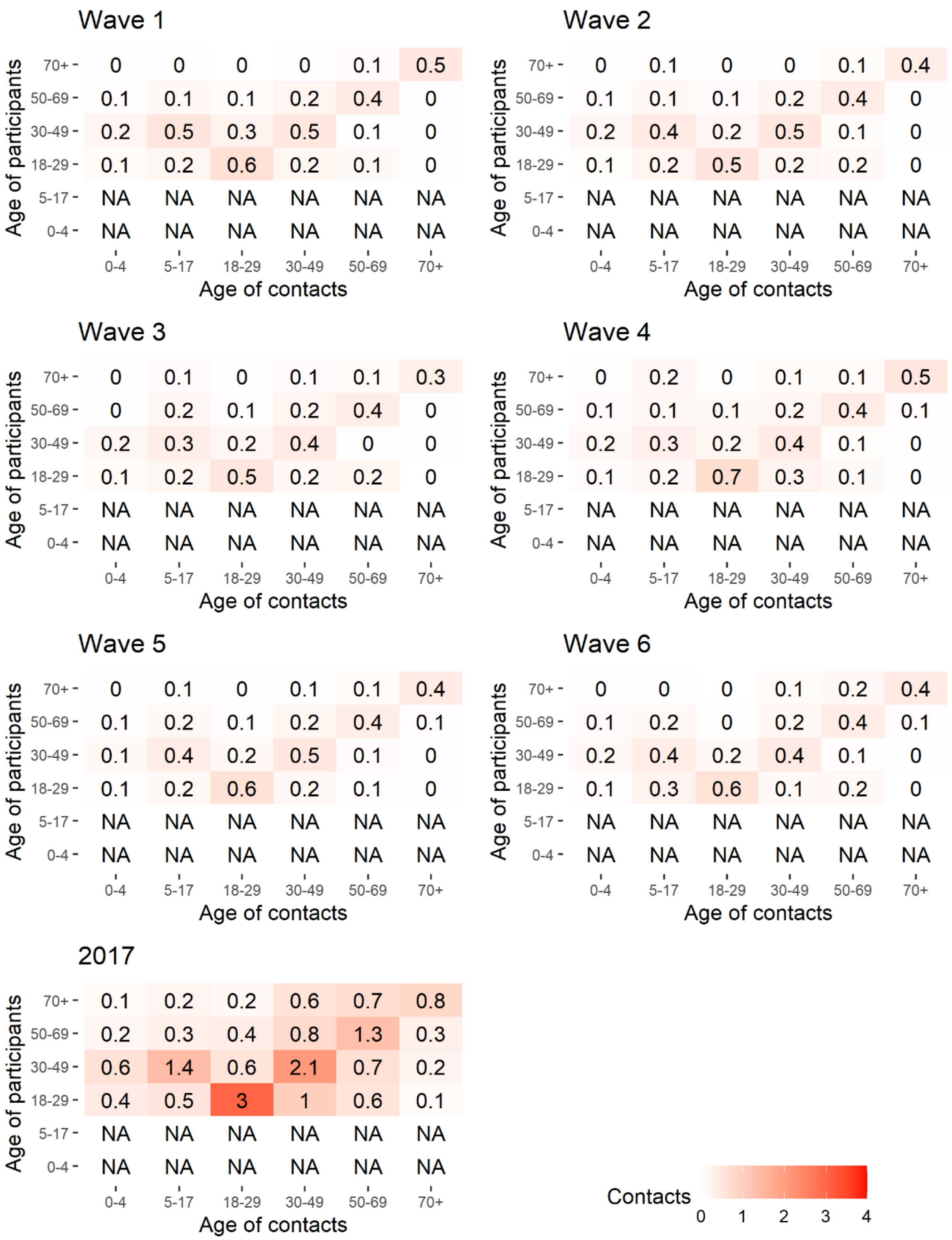

**Fig S8**: Imputed, setting-specific social contact matrices showing the mean number of daily physical contact for the six CoMix waves. Locations include all contacts made in the home, at schools, at workplaces and other community contacts (transport, sport activities etc.) The figures represent bootstrap mean values of N=10 000. Data were weighted on age and adjusted for reciprocity of contacts.

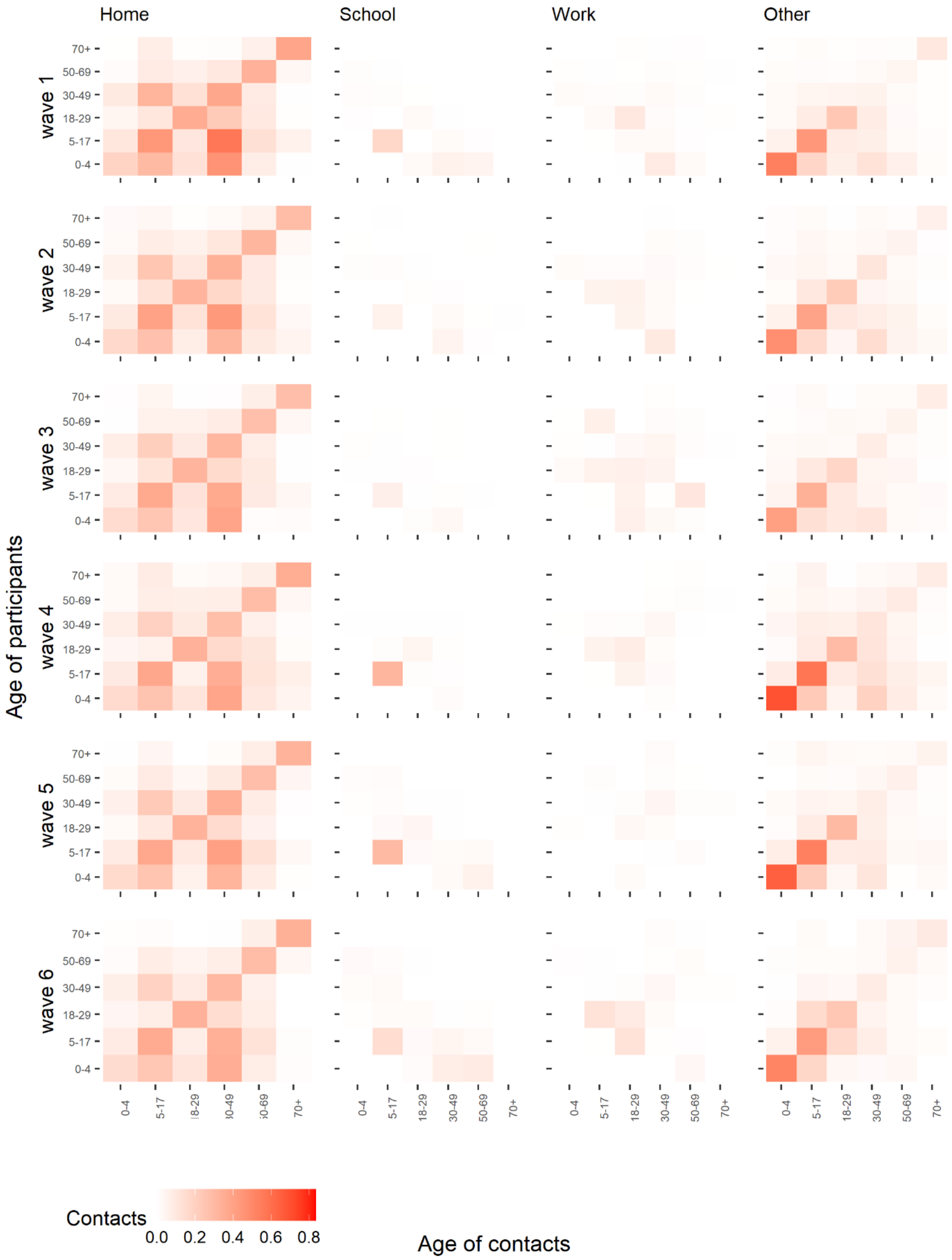

**Table S5:** Disassortativity measures from 2017 baseline survey and CoMix waves by type and location: standardized indices ($I_s^2$) with interquartile bootstrap intervals (IQR) of imputed matrices.

| | Location | 2017 Baseline ($I_s^2$) mean (Q1-Q3) | Wave 1 ($I_s^2$) mean (Q1-Q3) | Wave 2 ($I_s^2$) mean (Q1-Q3) | Wave 3 ($I_s^2$) mean (Q1-Q3) | Wave 4 ($I_s^2$) mean (Q1-Q3) | Wave 5 ($I_s^2$) mean (Q1-Q3) | Wave 6 ($I_s^2$) mean (Q1-Q3) |
|---|---|---|---|---|---|---|---|---|
| All | All | 0.43 (0.42-0.44) | 0.46 (0.46-0.47) | 0.55 (0.53-0.56) | 0.59 (0.57-0.61) | 0.53 (0.52-0.54) | 0.46 (0.44-0.47) | 0.48 (0.46-0.49) |
| | Home | 0.63 (0.61-0.65) | 0.52 (0.51-0.53) | 0.51 (0.50-0.52) | 0.48 (0.46-0.49) | 0.6 (0.58-0.61) | 0.49 (0.48-0.51) | 0.47 (0.45-0.52) |
| | School | 0.19 (0.18-0.20) | 0.47 (0.38-0.56) | 0.63 (0.27-0.88) | 0.31 (0.27-0.35) | 0.08 (0.06-0.09) | 0.17 (0.09-0.22) | 0.55 (0.31-0.74) |
| | Work | 0.36 (0.33-0.39) | 0.34 (0.32-0.35) | 0.52 (0.43-0.60) | 0.68 (0.54-0.80) | 0.32 (0.29-0.34) | 0.40 (0.35-0.42) | 0.38 (0.34-0.42) |
| | Other | 0.46 (0.45-0.48) | 0.47 (0.45-0.48) | 0.57 (0.53-0.60) | 0.51 (0.49-0.53) | 0.53 (0.50-0.55) | 0.47 (0.44-0.49) | 0.44 (0.42-0.44) |
| Physical | All | 0.50 (0.48-0.51) | 0.47 (0.46-0.48) | 0.50 (0.48-0.51) | 0.54 (0.51-0.56) | 0.55 (0.53-0.57) | 0.46 (0.44-0.48) | 0.51 (0.48-0.53) |
| | Home | 0.63 (0.6-0.65) | 0.57 (0.55-0.58) | 0.59 (0.57-0.60) | 0.57 (0.55-0.58) | 0.65 (0.63-0.67) | 0.56 (0.55-0.58) | 0.58 (0.56-0.60) |
| | School | 0.19 (0.17-0.21) | 0.67 (0.53-0.80) | 0.77 (0.67-0.87) | 0.44 (0.36-0.52) | 0.09 (0.06-0.11) | 0.55 (0.26-0.76) | 0.88 (0.50-1.17) |
| | Work | 0.33 (0.29-0.36) | 0.60 (0.52-0.67) | 0.38 (0.24-0.49) | 0.77 (0.43-1.03) | 0.22 (0.19-0.25) | 0.52 (0.34-0.66) | 0.35 (0.25-0.43) |
| | Other | 0.52 (0.5-0.55) | 0.43 (0.39-0.46) | 0.5 (0.47-0.57) | 0.46 (0.42-0.50) | 0.54 (0.50-0.58) | 0.45 (0.40-0.50) | 0.34 (0.30-0.38) |
